# The multivariate genetic architecture of psychiatric and insulin resistance multimorbidity

**DOI:** 10.1101/2024.10.02.24314704

**Authors:** Nina Roth Mota, Giuseppe Fanelli, Izel Erdogan, Marieke Klein, Emma Sprooten, Ábel Fóthi, I. Hyun Ruisch, Geert Poelmans, Kazi Asraful Alam, Jan Haavik, Theresa Wimberley, Martina Arenella, Alessandro Serretti, Chiara Fabbri, Barbara Franke, Janita Bralten

**Author notes:** Corresponding author*: Dr Janita Bralten, Radboud university medical center, Department of Human Genetics, P.O. Box 9101, 6500 HB Nijmegen The Netherlands. These authors contributed equally to this work (Nina Roth Mota, Giuseppe Fanelli).

## Abstract

Psychiatric disorders frequently co-occur with insulin resistance (IR)-related conditions, including obesity, type 2 diabetes mellitus (T2DM), and metabolic syndrome (MetS). Although pairwise genetic correlations have been observed, the shared genetics underlying this multimorbidity remains underexplored. Here, we investigate the joint genetic architecture of psychiatric-IR multimorbidity, explore tissue-specific gene expression associations, and identify potential underlying biological mechanisms and repurposable drugs. We applied genomic structural equation modeling (SEM) to genome-wide association study (GWAS) data (N=9,725–933,970) from five psychiatric disorders (attention-deficit/hyperactivity disorder, anorexia nervosa, major depressive disorder, obsessive-compulsive disorder, and schizophrenia) and three IR-related conditions (MetS, obesity, T2DM). Factor analyses revealed a 2-factor solution, where one of the factors was composed by all psychiatric disorders (excluding schizophrenia) and IR-related conditions (the Psych-IR factor), representing the shared genetics of these psychiatric and IR-conditions. This factor showed genetic correlations with the inferior temporal, lateral occipital, and total cortical brain surface areas. A multivariate GWAS of the Psych-IR factor identified 150 risk loci and 366 associated genes (128 novel). The significant gene-set associations included the insulin binding and the Notch signaling pathways, while the gene-property tissue expression implicated the cerebellum, brain cortex, and pituitary gland, particularly involving the brain during prenatal development stages. Transcriptome-wide SEM (T-SEM) assessed tissue-specific gene expression associations and identified 499 genes (191 novel), including MHC-related genes. Drug repurposing analysis using PharmOmics suggested six potential candidates, including memantine and rosiglitazone. Associated genes derived from the Psych-IR factor multivariate GWAS and T-SEM results were combined for enrichment analyses, which highlighted the involvement of the chr16p11.2 region, BDNF signaling, and lipid metabolism. The identified Psych-IR factor offers novel insights into the shared genetic and biological mechanisms underlying psychiatric-IR multimorbidity, providing a foundation for future research on precision medicine and prevention approaches.

## Introduction

The co-occurrence of psychiatric disorders and somatic insulin resistance (IR)-related conditions, such as obesity, type 2 diabetes mellitus (T2DM) and metabolic syndrome (MetS), is often observed (Perry et al., 2021; Wimberley et al., 2022). Population-based studies have demonstrated that obesity not only increases the risk of developing T2DM and metabolic syndrome but also elevates the likelihood of receiving a psychiatric diagnosis (Leutner et al., 2023). Moreover, large-scale Danish registry data reveal bidirectional associations between T2DM and various psychiatric disorders, including neurodevelopmental, mood, and psychotic disorders (Wimberley et al., 2022). This observed multimorbidity between IR-related conditions and psychiatric disorders complicates clinical trajectories (Kraus et al., 2023; Skou et al., 2022) and is linked to more severe clinical outcomes; for instance, T2DM has been associated to more severe depression and, conversely, depression is linked to higher rates of complications and mortality in T2DM (Fanelli and Serretti, 2022; Possidente et al., 2023).

Of note, IR generally refers to a reduced response to insulin stimulation on peripheral tissues, resulting in elevated blood glucose levels (DeFronzo et al., 2015; Gluvic et al., 2017). However, it is increasingly evident that insulin signaling disruption also has significant effects on the brain (Agrawal et al., 2021). Insulin receptors are expressed in most brain regions (Sullivan et al., 2023), and insulin is involved in important brain processes like synapse formation, neuroprotection, and neuronal survival (Pomytkin et al., 2018). A growing body of evidence links IR-related conditions with cognitive deficits across multiple domains (Fanelli et al., 2022b; Ottomana et al., 2023) and suggests that central IR affects key neurotransmitter systems, such as dopamine signaling, which is involved in reward-seeking behavior and cognitive function (Gruber et al., 2023). Additionally, IR affects brain structures that are part of the mesolimbic pathway (i.e., the ventral tegmental area and nucleus accumbens), as well as the hippocampus (Lyra E Silva et al., 2019), influencing both hedonic perceptions and cognitive functions (Fanelli and Serretti, 2022; Gruber et al., 2023). The prefrontal cortex is also susceptible to the effects of IR, which can result in impaired cognitive flexibility and working memory deficits (Arnold et al., 2018a; Willette et al., 2013). IR is also associated with brain regional atrophy in Alzheimer’s disease, particularly in the bilateral parietal-occipital junction and medial temporal regions, hippocampal and ventromedial prefrontal cortex volumes in bipolar depression and healthy subjects (Mansur et al., 2021; Morris et al., 2014; Mullins et al., 2017).

While many studies attribute metabolic disturbances in psychiatric patients to unhealthy lifestyles, sedentary habits, or the chronic use of psychotropic medications (e.g., (Grajales et al., 2019)), evidence suggests that these associations are not merely by-products of such factors. Glycemic and metabolic imbalances have been detected even in drug-naïve psychiatric patients at disorder onset, implying the potential involvement of shared pathogenic mechanisms (Garrido-Torres et al., 2021). Genetic studies reinforce the hypothesis of a shared biological basis for this multimorbidity showing significant genetic correlations between several psychiatric disorders—including attention-deficit/hyperactivity disorder (ADHD), anorexia nervosa (AN), obsessive-compulsive disorder (OCD), major depressive disorder (MDD), and schizophrenia (SCZ)—and IR-related conditions such as MetS, obesity, and T2DM (Fanelli et al., 2022a). Subsequent local genetic correlation analyses further demonstrated that these genetic overlaps are not always evenly distributed throughout the genome highlighting the complex genetic landscape of IR-neuropsychiatric multimorbidity ((Fanelli et al., 2024), medRxiv). Additionally, a family-based study indicated that relatives of individuals with a psychiatric disorder have an increased risk for T2DM (Wimberley et al., 2024). These findings suggest that shared underlying mechanisms are important for the multimorbidity between psychiatric disorders and IR-related conditions.

While bivariate genetic analyses have been instrumental for identifying shared genetic etiologies between pairs of psychiatric and IR-related conditions, the global joint genetic architecture and biological substrates underlying the multimorbidity across these two groups of conditions has not been explored. To address this gap, we employed genomic structural equation modeling (genomic SEM), a novel multivariate approach that enables analyzing the shared genetic architecture of multiple complex traits simultaneously (Grotzinger et al., 2019). This method allows for the identification of genetic variants associated with a common underlying genetic factor, shown to capture loci that are missed by traditional univariate genome-wide association study (GWAS) approaches (Grotzinger et al., 2019). Given that many genetic loci identified through GWAS likely exert their effects via modulation of gene expression (e.g., as expression quantitative trait loci or eQTLs; (Westra et al., 2013)), transcriptome-wide association studies (TWASs) can be helpful to quantify the effect of gene expression on complex traits (Gusev et al., 2016). Transcriptome-wide structural equation modeling (T-SEM) extends genomic SEM by modeling tissue-specific gene expression within a multivariate network of genetically overlapping traits, providing further insights into the molecular mechanisms involved (Grotzinger et al., 2022a). These transcriptomic results can also be integrated with open-source databases to identify potential, novel drug candidates (Y.-W. Chen et al., 2022).

In this study, we aimed to elucidate the joint genetic architecture underlying the multimorbidity of psychiatric disorders and somatic IR-related conditions. We applied genomic SEM to explore the genetic factor structure best explaining the shared genetics between five psychiatric disorders and three IR-related metabolic conditions that have previously shown significant pairwise genetic correlations (Fanelli et al., 2022a). Using the genomic SEM framework, we also examined the genetic relationships between the identified latent multimorbidity factor and brain morphometry (Grasby et al., 2020; Hibar et al., 2017; Satizabal et al., 2019), as well as estimated the effects of single-nucleotide polymorphisms (SNPs), genes, and gene sets on such a latent multimorbidity factor. Furthermore, employing T-SEM, we specifically investigated the association between brain-specific transcriptomic patterns and the identified multimorbidity factor, aiming to uncover genes whose tissue-specific gene expression might overlap with brain molecular signatures of repurposable drugs.

## Materials and methods

### Input univariate GWAS summary statistics

In order to explore the joint genetic architecture underlying the multimorbidity of psychiatric disorders and somatic IR-related conditions, we used GWAS summary statistics of European ancestry datasets of five psychiatric disorders (i.e., ADHD, AN, MDD, OCD, and SCZ) and three somatic IR-related conditions (i.e., MetS, obesity, and T2DM) that showed significant pairwise genetic correlations (Fanelli et al., 2022a) as input for the genomic factor analyses and further genomic SEM and T-SEM analyses (**Table 1**; see also **Figure 1a)**. SNP-based heritability was estimated using Linkage Disequilibrium Score Regression (LDSC; (Bulik-Sullivan et al., 2015)) and is reported on the liability scale. For details regarding sample ascertainment, phenotype description, quality control, and related procedures, we refer the reader to the corresponding univariate GWAS original publications listed on **Table 1**.

**Figure 1.**
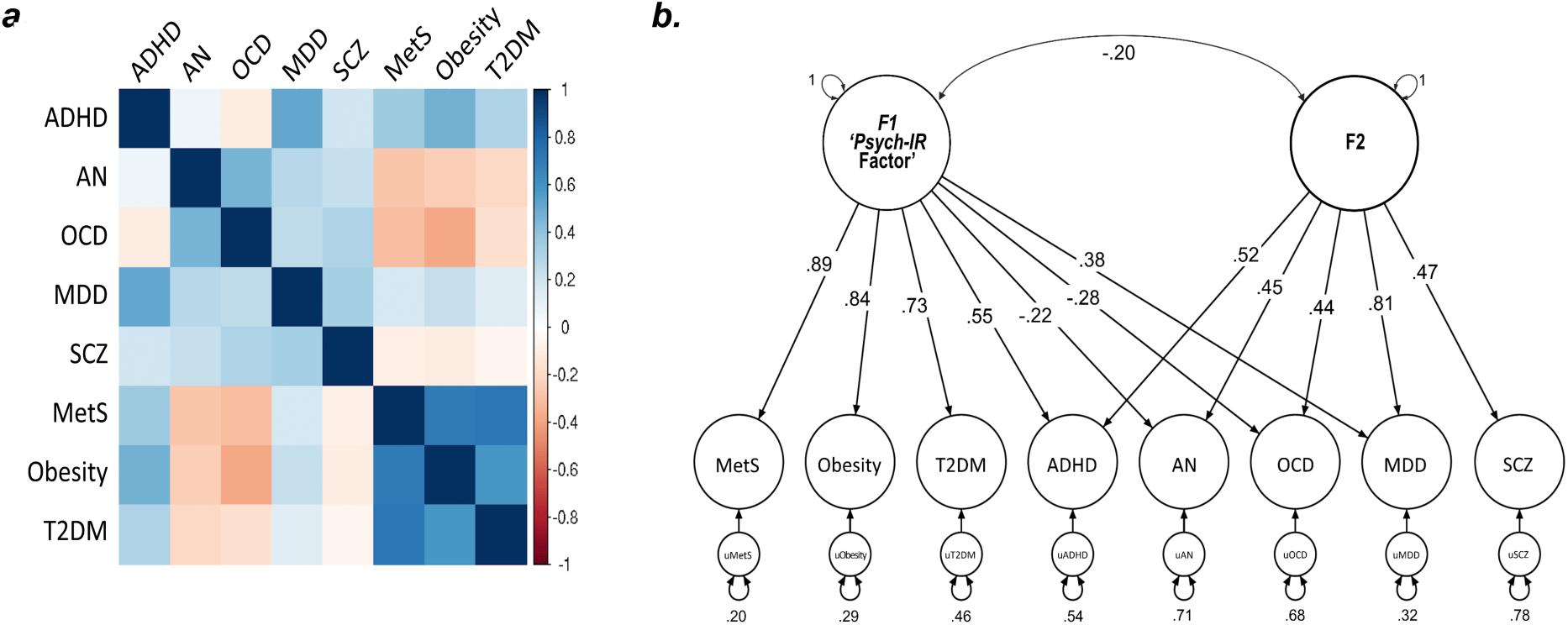
Multivariate genetic architecture of five psychiatric disorders and three somatic IR-related conditions. **a.** Heatmap of pairwise genetic correlations based on all autosomes estimated using LDSC regression within genomic SEM; **b.** Path diagram for the final confirmatory factor model with standardized parameter estimates. Circles represent the genetic components of each disorder, condition, or common genetic factor. One-headed arrows represent regression relationships from the independent variables pointing towards the dependent variables. Two-headed arrow between variables represent a covariance relationship. Two-headed arrows connecting the variable to itself represents residual variance. ADHD, attention-deficit/hyperactivity disorder; AN, anorexia nervosa; OCD, obsessive-compulsive disorder; MDD, major depressive disorder; SCZ, schizophrenia; MetS, metabolic syndrome; T2DM, type 2 diabetes mellitus.

**Table 1.** Contributing univariate genome-wide association study (GWAS) datasets.

| Univariate GWAS | Cases | Controls | Total sample | Population prevalence | SNP-based heritability (SE) | GWAS Reference |
| --- | --- | --- | --- | --- | --- | --- |
| ADHD | 38,691 | 186,843 | 225,534 | 0.087 | 0.213 (0.010) | (Demontis et al., 2023) |
| AN | 16,992 | 55,525 | 72,517 | 0.009 | 0.165 (0.011) | (Watson et al., 2019) |
| MDD | 170,756 | 329,443 | 500,199 | 0.21 | 0.290 (0.045) | (Howard et al., 2019) |
| OCD | 2,688 | 7,037 | 9,725 | 0.02 | 0.094 (0.004) | (IOCDF-GC/OCGAS, 2018) |
| SCZ | 53,386 | 77,258 | 130,644 | 0.01 | 0.223 (0.008) | (Trubetskoy et al., 2022) |
| MetS | 59,677 | 231,430 | 291,107 | 0.25 | 0.201 (0.011) | (Lind, 2019) |
| Obesity<sup>a</sup> | 9,805 | 235,085 | 244,890 | 0.39 | 0.267(0.025) | (Watanabe et al., 2019) |
| T2DM | 80,154 | 853,816 | 933,970 | 0.1 | 0.174 (0.008) | (Mahajan et al., 2022) |

Note. Population prevalences were used for the liability scale conversion and were retrieved from their original publications and/or (Grotzinger et al., 2019) for the psychiatric traits, from (O'Neill and O'Driscoll, 2015) for MetS, from (World Health Organization, 2018) for obesity, and from (Kumar et al., 2024) for T2DM.
Abbreviations: ADHD, attention-deficit/hyperactivity disorder; AN, anorexia nervosa; MDD, major depressive disorder; OCD, obsessive-compulsive disorder; SCZ, schizophrenia; MetS, metabolic syndrome; T2DM, type 2 diabetes mellitus; SE, standard error; GWAS, genome-wide association study. <sup>a</sup>GWAS ATLAS ID: 3687, UK Biobank phenotype field 41204 (41204\_E66), which refers to the trait 'Diagnoses - secondary ICD10: E66 Overweight and obesity'.

### Genomic Structural Equation Modeling

#### Genomic factor analyses

A multivariate extension of LDSC (Bulik-Sullivan et al., 2015) within genomic SEM; (Grotzinger et al., 2019)) was conducted in order to estimate genetic correlations between all pairwise combinations of the studied phenotypes (**Figure 1a**) and to generate three covariance matrix sets, which were based on the odd, even, or all autosomal chromosomes. Genomic SEM is not biased by sample overlap and is capable of accounting for differences in sample sizes among the univariate GWASs that are used as input. Standard procedures were followed and default filtering parameters for this munging step, such as retaining only SNPs that overlap with HapMap3 SNPs outside of the major histocompatibility complex (MHC) region and excluding SNPs with imputation quality (INFO)<0.9 and/or with minor allele frequency (MAF)<1%, were applied whenever such information was available for the univariate GWAS summary statistics. As basis for the multivariate LDSC, we used precalculated LD scores derived from the 1000 Genomes (Phase 3) European reference population (1000 Genomes Project Consortium et al., 2015; Bulik-Sullivan et al., 2015). The sample prevalence for all phenotypes was set to 0.5 in the LDSC estimation step since we used the effective number of samples as the sample size for the munge step, following the instructions provided in Genomic SEM GitHub page (<u>2.1 Calculating Sum of Effective Sample Size and Preparing GWAS Summary Statistics · GenomicSEM/GenomicSEM Wiki · GitHub</u>). The assigned population prevalence of each phenotype can be found in **Table 1**.

In order to model the genomic factor structure underlying the psychiatric and somatic IR-related conditions investigated here, we conducted a series of factor analyses based on the genetic covariance matrices derived from LDSC analyses within genomic SEM. We first conducted exploratory factor analyses (EFA) on the output of the LDSC analyzes with odd chromosomes using the *factanal* function of R with promax rotation, which allows factors to be correlated. We tested solutions up to three latent factors, while retaining factors that explained at least 20% of the variance. Based on the results of the EFA in odd chromosomes, we performed follow-up confirmatory factor analyses (CFA) for the one-factor and two-factor models using the genetic covariance matrix from the LDSC with even chromosomes, where factors were assigned to traits when their standardized loading exceeded .20 in the corresponding EFA. The model uses Diagonally Weighted Least Square (DWLS) and was specified so that the variance of each latent factor is fixed to 1 (i.e., unit variance identification).

Model fit was assessed using standard measures in structural equation modeling, as described in Grotzinger et al. (2019), where values >0.9/0.95 for the comparative fit index (CFI) and <0.10/0.05 for the standardized root mean square residual (SRMR) were considered reflective of an acceptable/good fit model. The Akaike Information Criterion (AIC) is a relative fit index, which can be used to compare models (i.e., lower AIC values indicating better fit). Chi-square p-values are often significant in genomic SEM analyses due to the high power of current GWASs; however, chi-square estimates may still be informative for comparing competing models (i.e., lower chi-square values indicating better fit). Finally, the CFA model with best fit in even chromosomes was also assessed for all autosomes.

#### Genetic correlation with brain morphometry

We used genomic SEM to assess the genetic link between the identified latent multimorbidity factor(s) and brain morphological traits. More specifically, we modeled the genetic covariances and correlations between the factor and the GWAS summary statistics of 1) the bilateral averages of cortical thickness and surface area (SA) of 34 brain regions and the total brain (N=33,992; (Grasby et al., 2020)); and 2) eight subcortical volumes (Hibar et al., 2017; Satizabal et al., 2019), namely nucleus accumbens (N=32,562), amygdala (N=34,431), brainstem (N=28,809), caudate nucleus (N=37,741), globus pallidus (N=34,413), putamen (N=37,571), thalamus (N=34,464), and hippocampus (N=33,536). All brain morphometry-related GWAS summary statistics underwent standard filtering and processing through the munge function of LDSC in genomic SEM, as detailed above. We refer the reader to the original publications ((Grasby et al., 2020), for cortical thickness and SA; (Satizabal et al., 2019) and (Hibar et al., 2017) for the eight subcortical volumes) for details about how these brain-related univariate GWAS were performed. Bonferroni correction was applied to account for multiple comparisons, thus adjusting the significance threshold (αBonf=0.05/78 brain phenotypes=6.41×10^-4^).

We further computed heterogeneity statistics (Qtrait) for the associations of the latent factor with the brain morphological traits, as described in (Grotzinger et al., 2022b). For each brain phenotype, the Qtrait heterogeneity index evaluates to which extent that trait operates through the latent factor. This is done by comparing a model in which the brain trait predicted the factor only to one in which it predicted the individual disorders/conditions that compose the latent factor. A significant Qtrait (P<6.41×10^-4^) indicates that the pattern of associations between the brain trait and the individual disorders/conditions is not well accounted for by the factor.

#### Multivariate GWAS of the multimorbidity factor

After identifying the CFA that best explained the observed genetic covariances among the psychiatric disorders and the IR-related somatic conditions, we used genomic SEM (Grotzinger et al., 2019) to conduct a multivariate GWAS, estimating individual SNP effects on the identified latent multimorbidity factor. Quality control procedures of the univariate GWAS summary statistics were performed following genomic SEM guidelines, which included restricting to SNPs with an INFO score >0.6 (when available) and to SNPs with MAF >1% in the 1000 Genomes phase 3 European reference panel (1000 Genomes Project Consortium et al., 2015). Only genetic variants present in all input univariate GWAS summary statistics were used. For this step, we used unit loading identification to scale the latent factor(s) (instead of unit variance identification used in the CFA), which also allows deriving the effective number of samples (Neff) for the latent factor(s) (Neff was estimated as described in Mallard et al).

Similarly to the Qtrait statistics described above, we also performed SNP-level tests of heterogeneity (QSNP) to evaluate whether each SNP had consistent pleiotropic effects on the factor components (i.e., input disorders/conditions) that effectively only operate via the shared liability (null hypothesis) or whether there was evidence of heterogeneity, indicating that the SNP effect is not fully mediated by the factor (Grotzinger et al., 2019).

### Gene, gene-set, and gene-property analyses of the Psych-IR multivariate GWAS results

The results of the multivariate GWAS of the multimorbidity latent factor were submitted to Functional Mapping and Annotation of Genome-Wide Association Studies (FUMA; version v1.5.6; (Watanabe et al., 2017)), using default parameters (if not otherwise specified). We used the FUMA SNP2GENE module to identify independent genomic risk loci, and independent genome-wide significant SNPs within each locus, employing the standard clumping algorithm (Watanabe et al., 2017). After the removal of all significant QSNPs (P< 5×10-8), as well as any SNP in LD with those (r2>0.1, 250Kb), from the multivariate GWAS summary statistics, this module was also used to implement Multi-marker Analysis of Genomic Annotation (MAGMA; v.1.08; (De Leeuw et al., 2015)) gene-based, gene-set, and gene-property (tissue expression) analyses. Gene-based p-values were computed for protein-coding genes by mapping SNPs located within genes according to Ensembl v110. MAGMA gene-set association analysis uses the complete gene-based results, (thus differing from enrichment analyses of prioritized genes, described below) to perform one-sided (positive) association tests for 17,023 gene sets from the Molecular Signatures Database (MSigDB v2023.1.Hs; (Liberzon et al., 2011)). Bonferroni correction was used to set the genome-wide significance threshold for the gene-based and gene-set analyses. MAGMA gene-property tissue expression analyses also use the gene-based results to test the associations with highly expressed genes in specific tissues, while conditioning on average expression across all tissue types. These tissue expression analyses were performed across 30 general tissues and 54 tissues types (GTEx v8; (The GTEx Consortium et al., 2020)), as well as 29 different ages of brain samples and 11 general developmental stages of the brain (BrainSpan; (Kang et al., 2011)) (for more detailed information, please see (Watanabe et al., 2017)). We also ran the FUMA analyses on the eight GWAS summary statistics of the individual phenotypes that served as input for genomic SEM, in order to compare the significant loci and genes identified for the multivariate GWAS. Genomic loci and genes associated with the latent factor that did not overlap with those associated with the individual phenotypes were considered as novel/unique to the multimorbidity factor.

### Transcriptome-wide structural equation modeling (T-SEM)

T-SEM (Grotzinger et al., 2022a) was employed to investigate the effects of tissue-specific gene expression on the multimorbidity factor representing the shared genetics of psychiatric disorders and insulin resistance (IR)-related conditions. This method enables the examination of tissue-specific gene expression within a multivariate model of genetically overlapping traits.

First, to ensure sufficient SNP-level overlap with the tissue-specific expression weights, the univariate GWAS summary statistics of the eight input phenotypes (**Table 1**) were reprocessed using the LDSC munging function, this time using the 1000 Genomes SNPs as reference (1000 Genomes Project Consortium et al., 2015) (as recommended by the developers guidelines for T-SEM; https://github.com/GenomicSEM/GenomicSEM/wiki/7.-Transcriptome-wide-SEM-(T-SEM)). The genetic and sampling covariance matrices of these munged summary statistics were estimated by multivariate LDSC as implemented in Genomic SEM and are used as input to T-SEM (Grotzinger et al., 2019).

Univariate, summary-based TWASs were then performed using FUSION (Gusev et al., 2016) to test the association between predicted tissue-specific gene expression and each individual trait. This association was estimated as a weighted linear combination of GWAS Z-statistics using pre-compiled functional weights from external reference datasets containing both tissue-specific gene expression and genotype data. In particular, we used 15 tissue-specific functional weight datasets, including 13 referring to brain tissues (i.e., amygdala, anterior cingulate cortex, caudate, cerebellar hemisphere, cerebellum, cortex, frontal cortex, hippocampus, hypothalamus, nucleus accumbens, putamen, cervical spinal cord C1, substantia nigra) and one to the pituitary gland from the GTEx v8 (The GTEx Consortium et al., 2020), as well as one referring to the brain prefrontal cortex from PsychENCODE (Gandal et al., 2018). The selection of pituitary and brain tissues for these analyses was supported by the tissue specificity of genes from the multivariate GWAS of the multimorbidity factor (described above).

The tissue-specific gene expression estimates for each gene produced by univariate TWASs were used to expand both the genetic covariance and sampling covariance estimated previously. Specifically, the *read_fusion* function in Genomic SEM was employed to standardize the gene expression estimates relative to the phenotypic variance, thus integrating them into the LDSC genetic covariance matrices. We then applied the *userGWAS* function to evaluate the effect of gene expression on the previously identified factor representing the shared genetic liability across psychiatric disorders and IR-related conditions.

Lastly, T-SEM was used to examine the associations of tissue-specific gene expression with the multimorbidity latent factor. We applied a Bonferroni correction to adjust for multiple testing across 16,542 unique genes, resulting in a significance threshold of αBonf=3.02×10^-6^. To identify genes with potentially trait-specific effects, we also computed gene heterogeneity statistics (QGene) as a chi-square difference test between a common pathways model (where gene expression predicts the multimorbidity latent factor) and an independent pathways model (where the gene expression only predicts specific psychiatric or IR-conditions defining the factor) (Grotzinger et al., 2022a). To ensure robustness, we excluded from the list of significantly associated genes those with significant QGene values using the same Bonferroni corrected threshold.

The MHC region was excluded from follow-up analyses due to its highly complex LD structure, which may confound genetic association signals and inflate the number of false-positive findings (Miretti et al., 2005). However, we conducted parallel T-SEM T-SEM analyses both excluding and including the MHC region to provide a comprehensive assessment of its potential impact on the results, and findings from the both T-SEM analyses are presented to ensure transparency and completeness in reporting.

### Drug repurposing analysis

To identify potential therapeutic candidates for the psychiatric-IR multimorbidity, we used PharmOmics, a comprehensive online platform for drug repurposing (https://mergeomics.research.idre.ucla.edu/runpharmomics.php#; (Y.-W. Chen et al., 2022)). PharmOmics is a species- and tissue-specific drug signature database that leverages transcriptomic data to facilitate the identification of repurposable drugs by comparing user-provided gene signatures for a trait of interest (i.e., the multimorbidity factor, in our case) with a curated database of drug-induced gene expression profiles (Y.-W. Chen et al., 2022). The PharmOmics database integrates transcriptomic data from human, mouse, and rat studies across more than 20 tissues, compiling over 18,000 drug-induced gene signatures for 941 drugs and chemicals. This database was curated from multiple sources, including the Gene Expression Omnibus (GEO), ArrayExpress, TG-GATEs, and DrugMatrix repositories. For our analysis, we used the list of genes derived from significant tissue-specific gene expression associations from our T-SEM results as input into the PharmOmics platform (https://doi.org/10.1016/j.isci.2022.104052). These genes were classified into upregulated and downregulated groups based on their respective T-SEM Z scores and submitted separately to PharmOmics. Specifically, a gene-overlap analysis was conducted (Y.-W. Chen et al., 2022) to determine the degree of overlap between the input gene lists (upregulated and downregulated genes) and the drug-induced gene signatures in the database. This analysis included calculating odds ratios to quantify the strength of association between the list of genes resulting from T-SEM and drug-specific gene expression signatures in the PharmOmics database. Fisher’s exact tests were used to assess the statistical significance of these overlaps, determining the likelihood that the observed overlaps occurred by chance. A signed Jaccard score was employed to evaluate the direction of the overlap between the gene sets. A positive signed Jaccard score indicates that the drug and T-SEM gene sets overlap with congruent expression changes (e.g., both upregulated or both downregulated), while a negative signed Jaccard score suggests that the drug and T-SEM gene set overlap with opposite expression changes (e.g., one upregulated and the other downregulated). The therapeutic relevance depends on the direction of the gene regulation and the desired therapeutic objective. For example, if a pathway is upregulated in psychiatric-IR multimorbidity, a drug that induces a negative signed Jaccard score (indicating an opposite regulation of the overlapping genes) may be of therapeutic interest to counteract the disease-related up-/down-regulation.

We selected drug repurposing candidates based on the following stringent criteria: 1) individual pharmacological molecules already approved by the Food and Drug Administration (https://www.accessdata.fda.gov/) for conditions other than psychiatric disorders; 2) those with evidence of blood-brain barrier permeability (ADMET features from https://www.drugbank.com/; https://github.com/12rajnish/DeePred-BBB); 3) drugs with available molecular signatures derived from nervous tissues in the PharmOmics database; 4) candidates showing consistent Jaccard scores and P-value significance across species, ensuring cross-species concordance and eliminating discordant effects; and 5) candidates with significant P-values and negative Jaccard scores, indicating an opposing gene regulation pattern that could potentially reverse disease-related molecular changes.

### Enrichment analyses of prioritized genes

The significantly associated genes identified by the MAGMA gene-based analysis of the multivariate GWAS were combined with genes whose tissue-specific expression was associated with the genomic latent multimorbidity factor in T-SEM analysis to compose a list of prioritized genes. This combined list of genes was used as input for the GENE2FUNC module in FUMA (version v1.5.6; 10.1038/s41467-017-01261-5) to conduct enrichment analyses to test for overrepresentation of the prioritized genes in pre-defined gene sets from the MsigDB (v2023;(Liberzon et al., 2011)), which include hallmark gene sets (MsigDB h), positional gene sets (MsigDB c1), curated gene sets (MsigDB c2), regulatory target gene sets (MsigDB c3), computational gene sets (MsigDB c4), ontology gene sets (MsigDB c5), oncogenic signature gene sets (MsigDB c6), immunologic signature gene sets (MsigDB c7) and cell type signature gene sets (MsigDB c8), as well as sets of reported genes from the GWAS-catalog (MacArthur et al., 2017). For the list of all gene sets tested, please see (https://fuma.ctglab.nl/tutorial#gene2func). Genes located within the MHC region were excluded from the analyses due to the extensive high linkage disequilibrium pattern in the region and hypergeometric tests were used for these evaluations. The background gene set, against which the prioritized genes were tested, consisted of all other (i.e., non-prioritized) protein-coding genes and the option of excluding the MHC region in FUMA was selected. Multiple testing correction was performed using the Benjamini–Hochberg (FDR) method by default, with corrections applied per data category or subcategory (e.g., hallmark genes, positional genes, different subcategories of curated gene sets, and so on). FUMA reported gene sets with an adjusted PFDR<0.05 and where the number of prioritized genes overlapping with the gene set was greater than two.

## Results

### Genetic factor structure underlying psychiatric and somatic IR-related conditions

We formally modeled the genetic covariance structure of the five psychiatric (ADHD, AN, MDD, OCD, and SCZ) and three somatic IR-related phenotypes (MetS, obesity, and T2DM) which are genetically correlated ((Fanelli et al., 2022); and **Figure 1a**). Descriptives of the input data can be found in **Table 1**. Exploratory factor analyses suggested the two-factor solution as the best model (variance explained: R^2^(F1)=32.1%, R^2^(F2)=20%, R^2^(Total)=52.1%), since the one-factor solution explained only 31.4% of the variance, while the third factor in a three-factor model explained only 15.2% of the variance and was not retained (**Supplementary Table S1**). Confirmatory factor analyses, both in the even chromosomes as in the full set of autosomes, confirmed that a two correlated factors model fits the data well (for all autosomes: χ^2^=78.559, df=15, Pχ^2^=1.28×10-10, AIC=120.559, CFI=0.978, SRMR=0.053; **Supplementary Table S2;** see also **Supplementary Table S3**) and revealed a small negative genetic correlation between the two factors (rg=-0.204; SE=0.043; P=2.02×10-6; **Figure 1b**). The first factor consists of all psychiatric disorders, except schizophrenia, and all somatic IR-related conditions investigated. This factor is hereafter referred to as the psychiatric and IR-related (Psych-IR) multimorbidity factor. The second factor consists of all five psychiatric disorders investigated, but none of the somatic ones.

Given our aim of unraveling the genetic architecture underlying the psychiatric and IR-related multimorbidity, subsequent results are focused on the Psych-IR multimorbidity factor, which was taken forward to investigate its relationship with brain morphometry, to conduct a multivariate GWAS, exploring it at multiple levels, as well as to conduct T-SEM and drug repurposing analysis.

### Genetic overlap between the Psych-IR multimorbidity factor and brain morphometry

We examined patterns of genomic correlations between the Psych-IR multimorbidity factor and brain morphometry (**Supplementary Figure S1**). We observed significant negative genetic correlations between the Psych-IR multimorbidity factor and total SA (rg=-0.151; SE=0.033; P=4.89×10^-6^) and inferior temporal SA (rg=-0.183; SE=0.045; P=4.60×10^-5^), while the factor had a positive genetic correlation with lateral occipital SA (rg=0.113; SE=0.032; P=5.01×10^-4^) (**Figure 2**). Follow-up Qtrait analyses were conducted to examine whether the genetic associations between the brain traits and the disorders/conditions are well accounted for by the identified latent factor. Qtrait index analyses revealed no significant sign of heterogeneity involving the three significant genetically correlated brain traits, indicating that the implication of these brain structures are indeed via the common pathway of the Psych-IR multimorbidity factor (rather than independent pathways of individual psychiatric disorders and somatic IR-related conditions). **Supplementary Table S4** provides genetic correlation estimates and Qtrait results for all brain traits analyzed.

**Figure 2.**
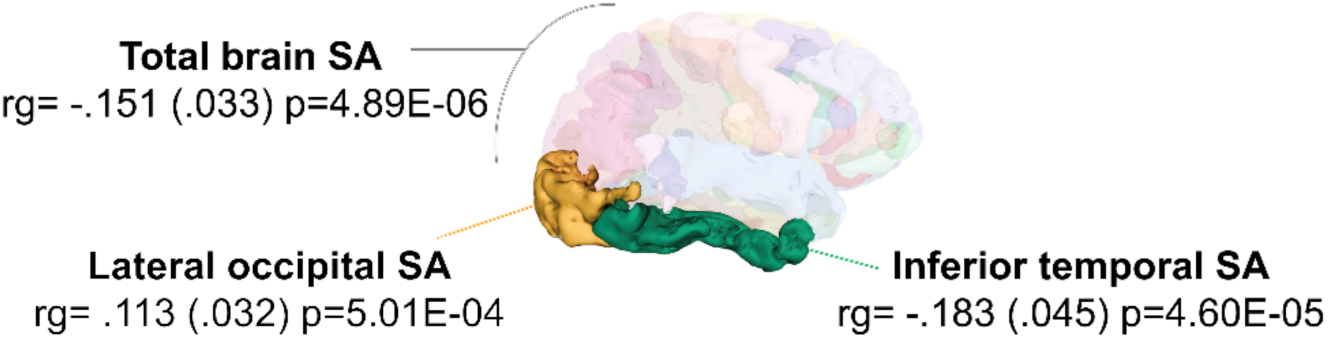
Genetic correlations (rg) between the Psych-IR multimorbidity factor and brain morphometric traits. Areas highlighted indicate the significant genetic correlations with total brain surface area (SA), lateral occipital SA, and inferior temporal SA. Visualization was performed using the ENIGMA-Vis tool (Shatokhina et al., 2021).

### Multivariate GWAS of the Psych-IR multimorbidity factor

Through a multivariate GWAS of the Psych-IR multimorbidity factor (Neff=622,007.6), we identified 11,672 genome-wide significant SNPs, which were distributed across 168 independent risk loci (**Supplementary Table S5**). We also performed QSNP heterogeneity tests in order to identify SNPs that act not through a common multimorbidity factor of psychiatric and IR-related somatic conditions, but directly on one or more of its components. There were 9,324 significant QSNPs (of which, 2,539 were genome-wide significant SNPs for the Psych-IR factor), indicating that the effects of these SNPs are not fully mediated by the latent genomic factor. Since we are interested in understanding the shared genetic basis of this multimorbidity, significant QSNPs were removed from downstream analyses (in order to reduce heterogeneity), along with those in LD (r^2^>0.1, 250Kb) with them. The final Psych-IR factor multivariate GWAS summary statistics contains 8,834 genome-wide significant SNPs, distributed across 150 independent loci (**Figure 3; Supplementary Figure S2**). Out of the 150 independent genomic loci identified, 46 of them did not overlap with the genomic loci associated in the input univariate GWAS of the psychiatric and somatic IR-related conditions that compose the latent factor (**Supplementary Table S6**).

**Figure 3.**
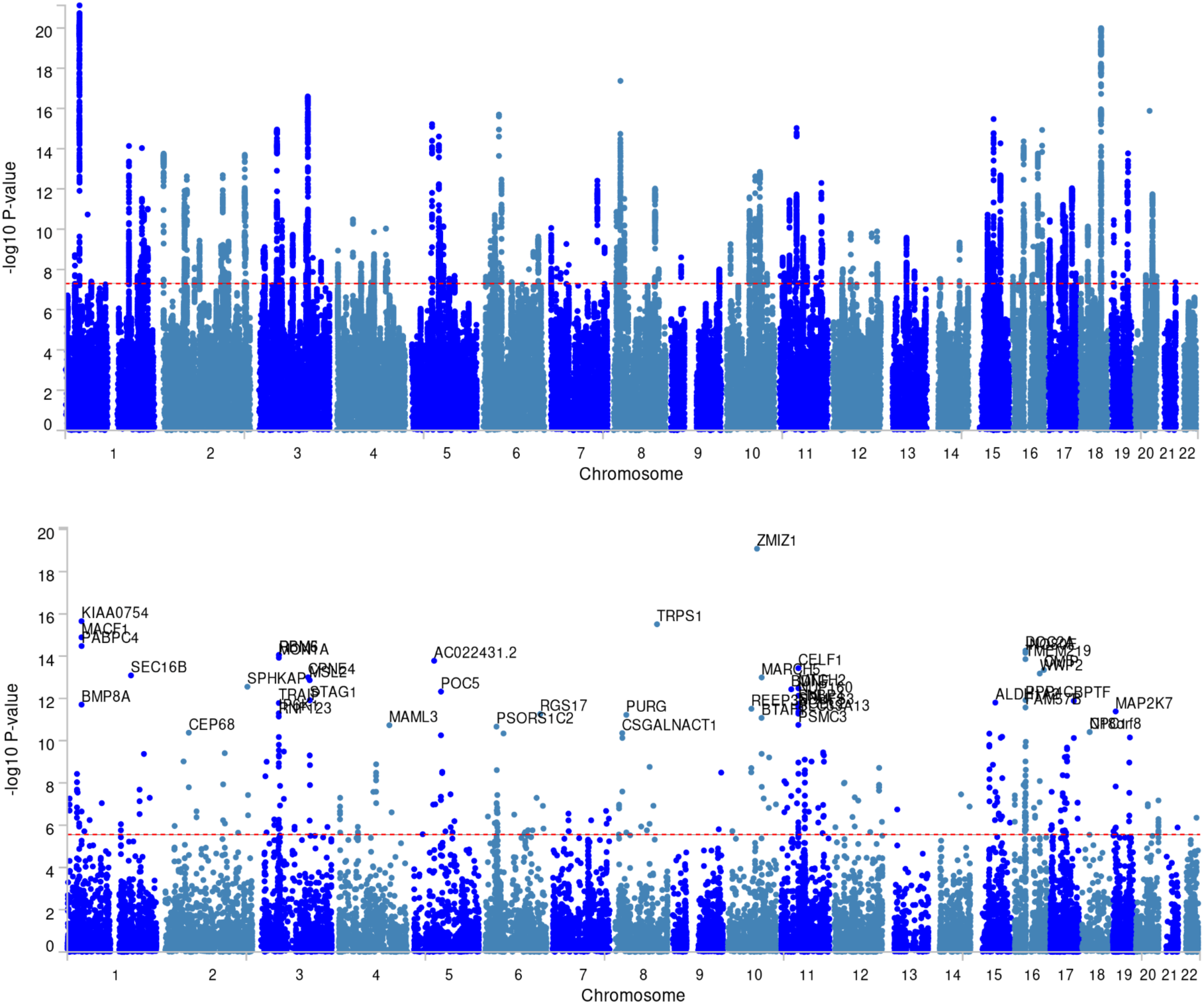
SNP-based (top) and gene-based (bottom) Manhattan plots of the Psych-IR multimorbidity factor multivariate GWAS. Dotted red line represents the genome-wide significance threshold of 5×10^-8^ for the SNP-based plot and of 2.78×10^-6^ (i.e., αBonf=0.05/17,977 protein coding genes) for the gene-based plot. Gene names are shown for the top 50 out of the 366 genome-wide significant genes. For a full list of significant genomic loci and associated genes, see **Supplementary Tables S6** and **S7**.

### Genes, gene sets and gene-property associations with the Psych-IR multimorbidity factor

Gene-based analysis identified 366 genome-wide associated genes (**Figure 3**). About one third of the associated genes (N=128) are considered novel, in the sense that they were not significantly associated with the individual phenotypes that compose the factor (i.e., genes were not significant in the individual input GWAS) (**Supplementary Table S7**). Gene-set analyses revealed six gene sets associated with the Psych-IR multimorbidity factor after Bonferroni correction for multiple testing, including one representing insulin binding (GOMF_INSULIN_BINDING; MsigDB M26667) and one implicating NOTCH signaling (REACTOME_SIGNALING_BY_NOTCH; MsigDB M10189), in addition to four gene sets of general Gene Ontology (GO) Biological Processes (**Table 2**).

**Table 2.** Gene sets significantly associated with the Psych-IR multimorbidity factor.

| Significant gene sets | N<sub>genes</sub> | P | P<sub>Bonf</sub> |
| --- | --- | --- | --- |
| GOBP_POSITIVE_REGULATION_OF_RNA_METABOLIC_PROCESS | 1,657 | 1.38x10<sup>-7</sup> | 0.0023 |
| GOBP_NEGATIVE_REGULATION_OF_BIOSYNTHETIC_PROCESS | 1,390 | 1.89x10<sup>-7</sup> | 0.0032 |
| GOBP_POSITIVE_REGULATION_OF_MACROMOLECULE_BIOSYNTHETIC_PROCESS | 1,723 | 5.48x10<sup>-7</sup> | 0.0093 |
| REACTOME_SIGNALING_BY_NOTCH | 183 | 7.60x10<sup>-7</sup> | 0.0129 |
| GOBP_NEGATIVE_REGULATION_OF_NUCLEOBASE_CONTAINING_COMPOUND_METABOLIC_PROCESS | 1,316 | 9.03x10<sup>-7</sup> | 0.0153 |
| GOMF_INSULIN_BINDING | 5 | 1.58x10<sup>-6</sup> | 0.0268 |

Abbreviations: N<sub>genes</sub>, number of genes included in the gene set; P, MAGMA gene-set association P-value; P<sub>Bonf</sub>, P-value after Bonferroni multiple testing correction for all the MsigDB gene sets tested.

Furthermore, MAGMA gene-property tissue expression analyses were performed to identify tissue specificity of the gene-based associations of the Psych-IR multimorbidity factor. Upon testing the relationships between the Psych-IR gene-based association results and tissue specific gene expression profiles, there were associations with the brain and pituitary general tissues types (**Supplementary Figure S3a**). A more fine-grained examination of the tissue types in question revealed that the Psych-IR factor was associated with highly expressed genes in specific brain tissues, namely the cerebellum, cerebellar hemisphere, cortex, and frontal cortex Brodmann Area (BA) 9, as well as the pituitary gland (**Supplementary Figure S3b**). The tissue expression analyses across 11 different general developmental stages of the brain implicated early, early-mid, and late-mid-prenatal stages (**Supplementary Figure S3c**), while no associations were found across the brain samples representing 29 different ages of the brain (BrainSpan; (Kang et al., 2011)).

### Multivariate TWAS

After excluding the MHC region and removing 31 unique genes (spanning 73 different gene-tissue pairs; **Supplementary Table S9**) with significant QGene values, T-SEM identified 462 unique genes whose expressions in the brain were associated with the Psych-IR multimorbidity factor (a heatmap of the most significant genes in each tissue and across tissues is depicted in **Supplementary Figure S4** and **S5,** respectively; **Supplementary Tables S10**). Among these, 188 were novel and not significant in any of the univariate TWASs of the input phenotypes (**Supplementary Tables S11**). Among the top significant up-regulated genes, *MST1R*, *MTCH2*, *RNF123*, *RP11-69E11.4*, *SNF8*, and *BMP8A* were recurrent across several tissues (**Supplementary Table S11 and Figure S5;** see also a Miami plot of the analysis including the MHC region in **Figure 4**). These genes are implicated in various biological processes including cell survival, migration and activation of macrophages (*MST1R*); mitochondrial function, apoptosis regulation, and lipid homeostasis (*MTCH2*); vesicle-mediated transport and and protein ubiquitination (*SNF8, RNF123*), and energy balance regulation (*BMP8A*). Among the top significant down-regulated genes, *RBM6*, *INO80E*, *RPAP1*, *C18orf8*, *VPS11*, and *MAPK3* were recurrent across tissues (**Supplementary Table S11**). These genes are involved in post-transcriptional modification (*RBM6*); chromatin remodeling (*INO80E*); vesicular trafficking (*VPS11*); and signal transduction (*MAPK3*). Of note, seven genes — *ANKDD1B, C17orf58, CRHR1, JMY, MAPT, PAM, and POC5* — demonstrated significant associations with the multimorbidity factor but showed discordant expression effects across different brain tissues (**Supplementary Table S12**).

**Figure 4.**
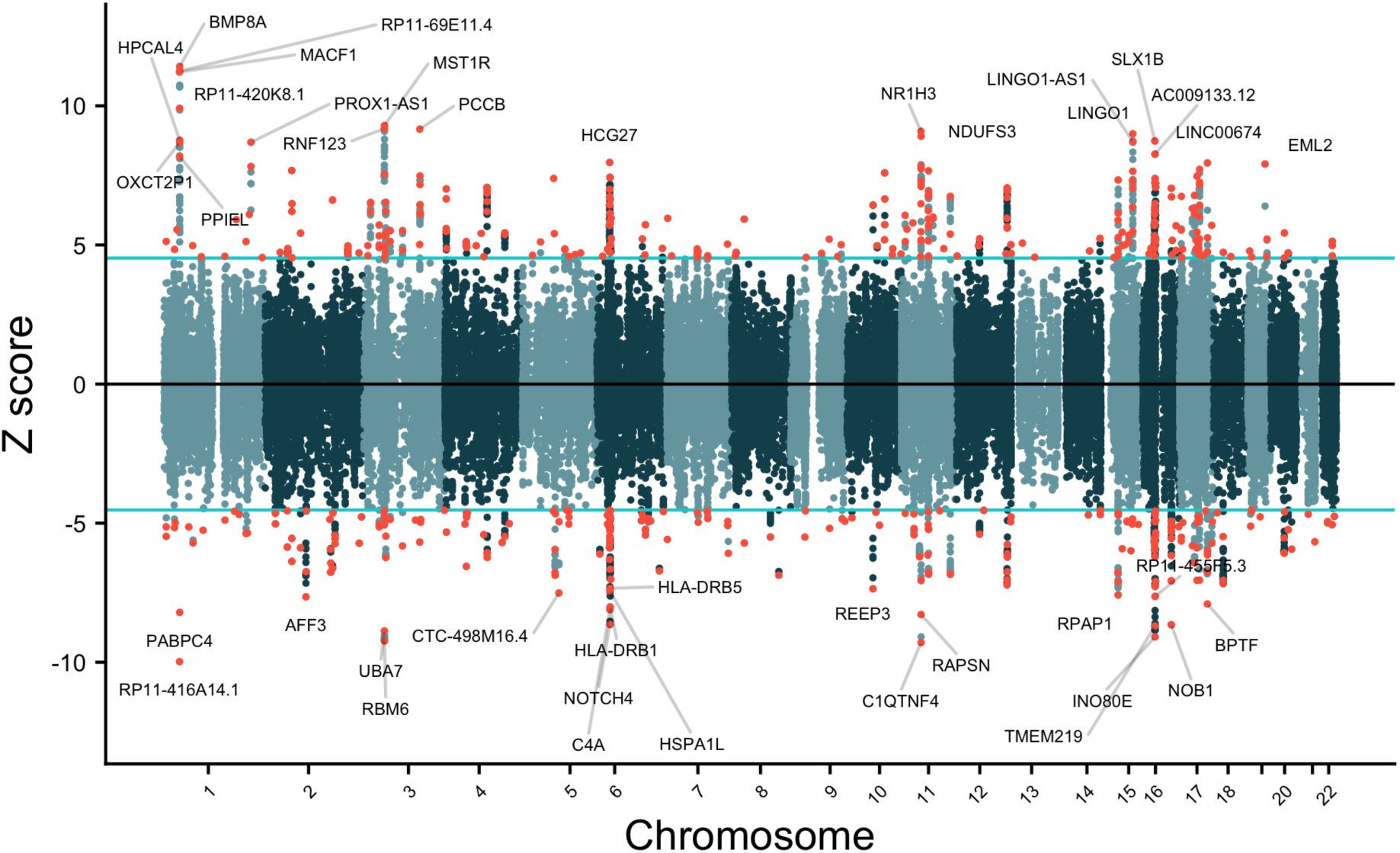
Miami plot of Z statistics for the estimated gene expression effects on the Psych-IR multimorbidity factor. Z statistics are signed such that orange dots on the upper and lower half of the plot reflect genes whose up-regulated and down-regulated expression, respectively, is significantly associated with the multimorbidity factor. The light blue horizontal lines reflect the Bonferroni-corrected significance threshold. Genes exceeding this threshold are shown as orange dots. For genes significant in multiple tissues, only the most significant instance is highlighted in orange. Up to 40 unique Bonferroni-significant genes are labeled across tissues. Genes having significant Q_Gene_ statistics were not included in this plot.

In the T-SEM analysis including the MHC region, 37 additional unique genes were significant (**Figure 4**; **Supplementary Figures S6-S7**), including three novel genes (i.e., *HSD17B8, RPS18, UQCC2*) that were not significant in any of the univariate TWASs (**Supplementary Table S13**). Among this region, the expression of the *HLA-DRB5* gene was the most frequently associated (significant across 14 tissues) with the multimorbidity factor, followed by *MICB* (12 tissues), and *CYP21A2* (11 tissues) (**Supplementary Table S14**). Among the up-regulated MHC genes, the most significant were *HCG27* in the brain anterior cingulate cortex, *CYP21A2* in the putamen, and *AGER* in the ACC. *HCG27* is a long non-coding RNA gene involved in various metabolic diseases; *CYP21A2*, a cytochrome P450 monooxygenase involved in mineralocorticoids and glucocorticoids biosynthesis; *AGER* plays a role in inflammatory responses and cellular signaling. Among the down-regulated MHC genes, the most significant were *NOTCH4* in the hippocampus and cerebellum, *C4A* in the cortex, and *HLA-DRB1* in the nucleus accumbens. *NOTCH4*, part of the Notch signaling pathway, is important for cell differentiation, proliferation, and apoptosis; *C4A*, a component of the classic complement pathway, is involved in immune responses and inflammation; *HLA-DRB1*, a major histocompatibility complex class II gene, is involved in antigen presentation and immune system functioning (**Supplementary Table S15**). These top three down-regulated genes were also the most significant ones among the whole set of MHC-related genes.

### Potential repurposable drugs

The overlap drug repurposing analysis using the PharmOmics platform identified six potential repurposable drugs for the Psych-IR multimorbidity factor (**Supplementary Table S16**). Among the evaluated compounds, bevacizumab emerged as a candidate in *Homo sapiens* (human) and demonstrated significant gene overlap in brain tissues. In *Mus musculus* (mouse), the analysis highlighted memantine, rosiglitazone, levodopa, cyclophosphamide, and ceftriaxone as leading candidates for repurposing. Memantine and rosiglitazone, known for their neuroprotective and anti-diabetic properties, respectively, showed robust overlap with the multimorbidity factor gene signatures. Additionally, levodopa, a precursor of dopamine and a commonly used drug in Parkinson’s disease, along with cyclophosphamide and ceftriaxone, both of which are involved in immune modulation and neuroprotection, also demonstrated significant overlap.

### Enrichment analyses of the Psych-IR prioritized genes

There were 534 protein-coding genes used as input for the combined gene set enrichment analyses, comprising 215 significantly associated genes derived from MAGMA gene-based analysis of the Psych-IR multivariate GWAS, 179 genes derived from the TSEM analyses, and 140 genes that were overlapping between these two approaches. There were 518 genes with unique Entrez IDs, which were compared against a total of 18,605 unique Entrez background protein-coding genes. After FDR correction for multiple testing within category/subcategory, we observed significant enrichments in 110 pre-computed sets from MsigDB (encompassing 104 unique gene sets) and in 201 sets of reported genes from the GWAS-Catalog (**Supplementary Table S8**). More specifically, these include the significant enrichments in 14 (MsigDB c1) positional gene sets. The most significant enrichment was in position chr16p11, which also had the highest proportion of overlapping genes with the gene set (i.e., 39 overlapping genes out of 97 in the gene set). There were also four significantly enriched (MsigDB c2) curated gene sets, two of which are also related with the chr16p11 region: the WP_16P112_PROXIMAL_DELETION_SYNDROME and the WP_16P112_DISTAL_DELETION_SYNDROME gene sets (both also significant in the analyses within the MsigDB c2:All Canonical Pathways (CP) and the MsigDB c2:CP/WikiPathways subcategories). Additional enrichments were found for three gene sets within the MsigDB c2:CP/WikiPathways subcategory, one of them related to the brain-derived neurotrophic factor (BDNF) signaling pathway and two related to familial hyperlipidemia; one MsigDB c5:GO:molecular functions gene set; three MsigDB c8 cell type signatures gene sets; and 79 MsigDB c3:Transcription Factor targets gene sets. No significant enrichment was observed among the MsigDB h, MsigDB c2:BioCarta, MsigDB c2:KEGG, MsigDB c2:Reactome, MsigDB c3:microRNA targets, MsigDB c4:All computational, MsigDB c4:Cancer gene neighborhoods, MsigDB c4:Cancer gene modules, MsigDB c5:GO:biological processes, MsigDB c5:GO:cellular components, MsigDB c6, and MsigDB c7 gene sets.

## Discussion

This study leverages state-of-the-art multivariate genomic and transcriptomic methods, including genomic SEM and T-SEM, to explore the genetic architecture underlying the frequent co-occurrence of psychiatric disorders and somatic IR-related conditions. We identified a latent Psych-IR multimorbidity factor representing the shared genomic liability across ADHD, AN, MDD, OCD, MetS, obesity, and T2DM, which provides novel insights into the biological underpinnings of their multimorbidity. The multivariate GWAS of the Psych-IR factor revealed 150 genomic loci and 366 associated genes, with many of these considered novel (i.e., not previously identified by the univariate GWASs that compose the factor). The insulin binding and the Notch signaling pathways were implicated with the Psych-IR factor. Genetic correlation analyses linked the Psych-IR multimorbidity factor to brain morphometry, including structures involved in visual and sensory processing. In addition, a series of tissue specificity analyses implicated specific brain areas, including the cerebellum, the brain cortex, and the pituitary gland. The integration of transcriptomic data by T-SEM revealed that the expression of 462 genes in the brain and pituitary gland is associated with the multimorbidity factor; these included 188 not previously detected in univariate TWASs. Top up-regulated genes, such as *MST1R*, *MTCH2*, and *BMP8A*, suggest roles for immune modulation, mitochondrial function, and energy balance, while down-regulated genes like *RBM6*, *INO80E*, and *MAPK3* highlight disruptions in chromatin remodeling and signal transduction.

Our findings advance the current understanding of the genetic underpinnings of psychiatric and IR-related multimorbidity, building upon previous studies that primarily explored pairwise correlations between psychiatric disorders and IR-related conditions (Fanelli et al., 2024, 2022a; Hübel et al., 2019), which, while informative, do not capture the joint genetic architecture underlying these multiple conditions. Our multivariate approach reveals that psychiatric disorders share common genetic variants and mechanisms with IR-related conditions, albeit with opposite loadings on the Psych-IR factor, highlighting the presence of a joint genetic architecture underlying the multimorbidity. Both the positive loadings for ADHD and MDD on the Psych-IR factor, as well as the negative loading of AN and OCD, are consistent with the direction of their pairwise genetic correlations (Fanelli et al., 2022a). The divergent pleiotropic effect observed with AN is also consistent with most clinical observations for AN, which is mainly characterized by weight loss, as opposed to the IR-related conditions (Walsh et al., 2023; American Psychiatric Association, 2013). While epidemiological data showed increased co-occurrence of OCD and T2DM (Wimberley et al., 2022), recent familial analyses indicate that parental T2DM was significantly less frequent in individuals with OCD, in line with negative genetic correlations, and indicating that phenotypic associations might be explained by other factors (like psychiatric comorbidities, shared environment or lifestyle factors) (Wimberley et al., 2024). Despite the well-documented clinical overlap between schizophrenia and metabolic dysregulation, particularly in the context of antipsychotic medication use, schizophrenia exhibited a weaker genetic loading and was ultimately not included in the Psych-IR multimorbidity factor. This may reflect the underlying genetic complexity given that previous local genetic correlation analyses indicate both positive and negative genetic local genetic correlations between schizophrenia and IR-related conditions (Fanelli et al., 2024). In addition, the metabolic side effects of antipsychotic medications used for treating schizophrenia include significant weight gain and IR, which are well-established but are likely driven by pharmacological mechanisms rather than by the genetic factors.

A key contribution of this study is the identification of genetic loci implicated in the psychiatric-IR multimorbidity, including novel genes that were not previously associated with individual psychiatric or IR-related phenotypes, while also reinforcing the involvement of established candidate biological pathways implicated in psychiatric-IR multimorbidity. In particular, among the top genes identified by the multivariate GWAS of the Psych-IR factor, the most significantly associated gene was *ZMIZ1*, which regulates transcription factors and interacts with nuclear hormone receptors. This gene shows genome-wide significant association also in the univariate T2DM GWAS and has recently been appointed as a novel regulator of brain development associated with ASD and intellectual disability (K. C. et al., 2024). *DOC2A*, located in the chr16p11 region, is involved with Ca^2+^-dependent neurotransmitter release and is mainly expressed in the brain. Other top genes are involved with interactions of cytoskeletal elements (e.g., *MACF1*), encoding transcription factors (e.g., *TRPS1*), mRNA stability (e.g., PABPC4), and tumor suppression (e.g., *RBM5, RBM6*). In terms of novel genes, the top three genes (*KCTD13, GDPD3, MAPK3*) are situated in the chromosome 16p11.2 region, discussed in more details below. These are followed by *MST1*, whose receptor, *MST1R*, was the top up-regulated gene in the T-SEM results and is directly involved in immune-inflammatory pathways (Huang et al., 2020). Among the other T-SEM top up-regulated genes across several tissues, *MTCH2* is involved in adipocyte differentiation and energy production (Peng et al., 2024). *RNF123/KPC1* and *SNF8* are linked to maintaining cellular homeostasis and regulating immunity (Kravtsova-Ivantsiv et al., 2015; Kumthip et al., 2017). Among the top down-regulated genes, *RBM6* and *INO80E* are involved in DNA repair and splicing/chromatin remodeling (Conaway and Conaway, 2009; Machour et al., 2021), pointing to disruptions in gene expression regulation. Among the novel genes, *STX4, EHD4*, and *USP46* participate in neurotransmission and insulin signaling, indicating a dual function in neuronal activity and glucose metabolism, and *ZNF268, MCM9* are involved in transcriptional regulation and genomic stability. Collectively, the novel genes highlight mechanisms that intersect both central nervous system function and peripheral metabolic regulation.

While analyses including the MHC region need to be interpreted cautiously given the genetic complexity due to the extensive LD, high gene density, and considerable allelic diversity of this region, they also highlighted immune-related genes as well. Among them, *HLA-DRB5* is involved in regulating immune responses and has been implicated in various brain-related and metabolic conditions, including SCZ, MDD, Parkinson’s disease, and both type 1 diabetes and T2DM (Ahmed et al., 2012; Jacobi et al., 2020; Santiago et al., 2023; Zhao et al., 2016). In addition, the *MICB* gene is a marker of cellular stress and tag cells for elimination triggering the activation of natural killer and CD8+ T cells (Derby et al., 1992) and its association might support the idea that cellular stress-induced immune dysregulation might be a common mechanism in psychiatric-IR multimorbidity. *CYP21A2* is involved in the biosynthesis of glucocorticoids and mineralocorticoids (Slominski et al., 2020). Glucocorticoids affect neuroplasticity and the expression of BDNF, essential for synaptic integrity and cognitive function (Tsimpolis et al., 2024). These findings related to the MHC region align with previous evidence highlighting key genes emerging from genetic annotations of loci correlated between psychiatric and IR-related conditions (Fanelli et al., 2024).

Our gene-set analysis on the Psych-IR multivariate GWAS results highlighted an association with the insulin binding and the Notch signaling pathways, reinforcing the hypothesis that metabolic dysregulation is central to the shared biological basis underlying the multimorbidity observed between psychiatric and IR-related conditions. The insulin binding gene set comprises five genes, three of which - *IDE*, *IGF1R*, and *INSR* - show genome-wide significant associations themselves in the gene-based analyses. *IDE* encodes the insulin-degrading enzyme which has been associated with T2DM, but also plays a role in cognitive processes and neurodegeneration (Henderson and Poirier, 2011) *INSR* encodes for the insulin receptor and insulin binding to this receptor activates pathways such as the PI3K-AKT/PKB pathway, responsible for most metabolic actions, and the Ras-MAPK pathway, which regulates gene expression and cell growth (Boucher et al., 2014). Dysregulation of these pathways has been implicated in both metabolic and neuropsychiatric outcomes, suggesting a shared mechanistic pathway (Borrie et al., 2017; Chen et al., 2024). *IGF1R* encodes the insulin-like growth factor 1 receptor, which is involved in neurogenesis, synaptic plasticity, and neuroprotective processes (Cardoso et al., 2021; Dyer et al., 2016). Although IGF1R’s role has been explored in relation to various cognitive functions (Cardoso et al., 2021), its specific link to the genetic architecture of psychiatric and metabolic comorbidity represents a novel finding in our study, as it was not identified as significant in any of the input univariate GWAS datasets. While the potential involvement of insulin signaling in psychiatric disorders is not a new concept (McIntyre et al., 2010), our findings clearly highlight the association of such a core insulin-related gene set with a genetic latent factor encompassing both conditions. This reinforces the need to explore this pathway further as a gateway for managing the co-occurrence of psychiatric disorders and somatic IR-related conditions.

Another gene set that showed significance to the Psych-IR factor was the Notch signaling pathway, which has also garnered attention for its potential role in both IR and the brain. Notch signaling is involved in the regulation of metabolic processes, particularly in the liver and adipose tissues. For instance, active Notch signaling correlates with IR and nonalcoholic fatty liver disease, indicating that Notch signaling may influence glucose metabolism through its effects on hepatic function (Valenti et al., 2013). Additionally, a mouse model overexpressing the Notch intracellular domain in adipocytes led to severe IR, thereby establishing a direct link between Notch signaling and metabolic dysregulation (Chartoumpekis et al., 2018). Noteworthy, Notch signaling was also involved in learning, memory, and social behavior, which are often disrupted in psychiatric disorders (Salazar et al., 2020), and it has also been implicated in neurodevelopment, neuronal connectivity and neurogenesis (Zhang et al., 2018), although a direct link with psychiatric disorders is currently missing (Salazar et al., 2020).

Subsequently, when combining the genome-wide significant genes from the Psych-IR multivariate GWAS with the associated genes from the T-SEM analyses, additional gene sets were implicated through the significant enrichment of our prioritized genes. Noteworthy are the ones related to proximal and distal chromosome 16p11.2 deletion syndrome, the BDNF signaling pathway, and the ones related to familial hyperlipidemia (types 3 and 4). Both the proximal and distal 16p11.2 deletion syndromes are rare genetic conditions caused by the deletion of around a 600kb and a 220 kb region, respectively, of chromosome 16 (OMIM#611913 and OMIM#613444, respectively). They are both characterized by symptoms related to both psychiatric and IR-related phenotypes, and mild intellectual disability and speech problems are also frequent among individuals with 16p11.2 deletion syndromes. Over 80% of the carriers of the proximal 16p11.2 deletion exhibit psychiatric disorders and obesity is a major comorbidity, affecting 50% of the carriers by age 7 and with a penetrance of 70% among adults (Zufferey et al., 2012). In a study comparing different 16p11.2 deletions, the vast majority of the individuals with proximal 16p11.2 deletion syndrome had developmental delays (85.5%), 19.4% autism spectrum disorder (ASD), 27.3% ADHD, 29.5% obesity, and 41% reported hyperphagia (Vos et al., 2024). In the same study, cases with distal 16p11.2 deletion showed the most severe obesity phenotype (73.7% obesity), with most cases presenting hyperphagia (61.1%), 40% intellectual disability, and 22.2% ASD (Vos et al., 2024). The enrichment of the BDNF signaling pathway also highlights a potential role of BDNF in bridging metabolic and psychiatric disorders. During development, the protein encoded by the *BDNF* gene promotes neuronal survival and differentiation and regulates synaptic plasticity, essential for adaptive neuronal responses, including long-term potentiation, and homeostatic regulation of excitability (Park and Poo, 2013; Rutherford et al., 1998). Its involvement in psychiatric conditions such as MDD, SCZ, and anxiety disorders is well-documented (Castrén and Kojima, 2017; Molendijk et al., 2014). Beyond its neural functions, BDNF plays a significant role in metabolic regulation. BDNF signaling intersects and shares downstream mechanisms with insulin pathways through its binding to tyrosine kinase B (TrkB) receptor (Bathina and Das, 2015). Moreover, low BDNF levels are associated with glucose impairment and lipid dysregulation, further implicating BDNF in metabolic health (Krabbe et al., 2007; Xia et al., 2022). This interaction is reinforced by findings that IR promotes neuroinflammation, which can impair BDNF signaling, creating a vicious cycle that exacerbates both metabolic and psychiatric conditions (Lima Giacobbo et al., 2019; Wei et al., 2021). Interventions such as exercise, which increase BDNF levels, have been shown to improve both insulin sensitivity and cognitive function (Dadkhah et al., 2023). Therefore, pharmacological strategies targeting BDNF signaling pathways could offer new avenues for treating metabolic and psychiatric disorders concurrently. We also observed significant enrichment of gene sets associated with familial hyperlipidemia types 3 and 4. Type 3 primarily involves impaired clearance of intermediate-density lipoproteins (IDL) due to mutations in the *APOE* gene, of which the protein plays a role in lipid transport and metabolism (Javvaji et al., 2024). *APOE* is also one of the Psych-IR genome-wide significant genes, and the most well-known risk gene for Alzheimer’s disease (Jackson et al., 2024). Familial hyperlipidemia type 4, or familial hypertriglyceridemia, involves increased levels of VLDL in the blood, driven by both enhanced production and decreased clearance (Goyal et al., 2024). Dyslipidemia is a common feature in both psychiatric conditions, such as MDD and SCZ, and somatic ones like MetS, where lipid abnormalities may exacerbate IR by promoting chronic inflammation, oxidative stress, and endothelial dysfunction (Higashi, 2023; Zorkina et al., 2024). The enrichment of these gene sets suggests a role for lipid metabolism in the pathophysiology of the multimorbidity of psychiatric and IR-related conditions.

In terms of brain morphometry implication, our analysis revealed significant negative genetic correlations between the Psych-IR multimorbidity factor and both total SA and inferior temporal SA. The inferior temporal cortex is primarily involved in visual processing, especially object and face recognition (Conway, 2018), as well as the retrieval of visual memories (Mruczek and Sheinberg, 2007). This region has been closely linked to metabolic dysfunctions, including obesity and IR (Morris et al., 2014; Opel et al., 2021). For instance, a Mendelian randomization study demonstrated that higher waist-hip ratio causally reduces the surface area of the inferior temporal cortex (Chen et al., 2023). In addition, positive genetic correlation was found for the Psych-IR multimorbidity factor and the lateral occipital cortex, which is involved in the perception of shapes and forms, as well as the processing of visual stimuli in a multisensory context (Zhang et al., 2004). Altered glucose metabolism in this region has been linked to cognitive impairments in various conditions, including SCZ and T2DM (Wijtenburg et al., 2019). Studies have demonstrated that hypoperfusion in the occipital regions, including the lateral occipital cortex, correlates with higher IR and deficits in visual memory performance, particularly in patients with T2DM (Cui et al., 2017). This aligns with findings that neuronal IR biomarkers are significantly associated with memory measures and brain glucose levels, particularly in visual processing areas like the lateral occipital cortex (Wijtenburg et al., 2019). Consistent with our findings, previous studies found that IR is associated with smaller cortical gray matter volume, but not with subcortical gray matter volume in individuals with MetS (Lu et al., 2021). Another link to the brain is found in the tissue expression specificity of the Psych-IR gene associations, where our findings reveal that the Psych-IR multimorbidity factor is significantly associated with genes highly expressed in the pituitary gland and brain tissues, implicating specifically the cerebellum/cerebellar hemisphere, and cortex/frontal cortex BA9. While the cerebellum and cerebellar hemisphere have traditionally been linked to motor control, recent studies increasingly recognize their roles in cognitive and emotional regulation, as evidenced by studies linking cerebellar dysfunction to various psychiatric conditions, including mood disorders (Adamaszek et al., 2017; Schmahmann, 2019). Recent evidence indicates that individuals with high IR exhibit reduced gray matter volume and altered functional connectivity in the cerebellum, suggesting that IR can lead to significant neuroanatomical and functional connectivity changes in this region (Chen et al., 2014; H.-Y. Zhang et al., 2024). IR also correlates with reduced glucose metabolism in the cerebellum and frontal regions (Y. Chen et al., 2022). The frontal cortex, particularly Brodmann area 9 (BA 9), plays a role in executive functions, including decision-making, working memory, and cognitive control (Friedman and Robbins, 2022; Miller and Cohen, 2001), all of which are processes heavily implicated in psychiatric disorders. Previous work indicates that insulin signaling is essential for maintaining synaptic plasticity and neuronal health in the frontal cortex, and disruptions in insulin signaling can impair cognitive functions linked to the frontal cortex (Arnold et al., 2018b; Fanelli et al., 2022b; Kleinridders et al., 2014). The observed association with gene expression in the pituitary gland might suggest a link to the hypothalamic-pituitary-adrenal (HPA) axis, which regulates both the stress response and metabolic function. Dysregulation of the HPA axis is a well-established factor in psychiatric disorders and metabolic conditions, and might indicate a shared pathway that influences both groups of phenotypes and their co-occurrence (Joseph and Golden, 2017; Stetler and Miller, 2011). Moreover, the association of gene expression with early, early-mid, and late-mid prenatal developmental stages suggests that the genetic factors underlying the Psych-IR factor may exert their effects during critical periods of brain development. This finding aligns with the hypothesis that prenatal or early-life factors can shape the long-term risk for both metabolic and psychiatric disorders (Edlow, 2017). Prenatal exposures, such as maternal stress, poor nutrition, or gestational diabetes, could interact with genetic predispositions to alter brain development, thereby increasing susceptibility to both psychiatric disorders and metabolic dysregulation in offspring (Van Lieshout et al., 2011).

From a clinical perspective, our results indicating a shared genetic etiology between multiple psychiatric and psychiatric and IR-related somatic conditions highlights the need for a holistic approach in medicine, integrating both worlds in clinical care. Through the genomic approaches addressed in this manuscript we identified potential drug repurposing candidates, including memantine, rosiglitazone, levodopa, cyclophosphamide, bevacizumab, and ceftriaxone, that could offer possibilities for developing targeted therapeutic strategies aimed at addressing both psychiatric symptoms and IR. Memantine, an NMDA receptor antagonist, has shown efficacy in improving cognitive and negative symptoms in SCZ, as well as in counteracting excessive glutamate neurotransmission and related neurotoxicity in Alzheimer’s disease (Czarnecka et al., 2021; Zheng et al., 2018), and rosiglitazone, a peroxisome proliferator-activated receptor gamma (PPAR-γ) agonist, enhances neuronal insulin receptor function and provides neuroprotective effects (McIntyre et al., 2007; Pipatpiboon et al., 2012). Cyclophosphamide, an immunosuppressive agent, shows promise in managing severe IR and autoimmune encephalitis, which often accompany psychiatric symptoms (Dinoto et al., 2022; Yang et al., 2017). Bevacizumab, an anti-VEGF monoclonal antibody, could enhance glucose uptake via the upregulation of glucose transporters in response to the induced hypoxia, and it has been shown to improve cognitive function in a Alzheimer’s disease animal models (Heijmen et al., 2014; Kuang et al., 2017; M. Zhang et al., 2024). Other drug repurposing candidates might need careful consideration, like levodopa, used for Parkinson’s disease management due to its potential to exacerbate IR and disrupt glucose regulation, particularly in patients with pre-existing metabolic conditions (Smith et al., 2004). Ceftriaxone, a third generation cephalosporin antibiotic, presents challenges due to its impact on gut microbiota, which can lead to dysbiosis and decreased short-chain fatty acid production, ultimately exacerbating IR (Holota et al., 2019; Miao et al., 2021). Future research might prioritize the most promising candidates, which could be considered for further investigation in randomized-controlled trials as potential therapies for psychiatric-IR multimorbidity.

The strengths of this study lie in the use of large-scale GWAS datasets, advanced genomic SEM techniques, and the integration of transcriptomic data, which collectively provide a robust and comprehensive analysis of the genetic underpinnings of psychiatric and IR-related multimorbidity. These approaches allowed us to identify shared genetic factors that may not be detectable through traditional, univariate GWAS/TWAS analyses, thereby offering novel insights into the genetic and biological bases of psychiatric-IR multimorbidity. However, this study also has some limitations. First, our understanding of the functions of the identified genes and their roles in molecular pathways remains incomplete. While the discovery of novel loci is promising, further research is needed to elucidate their precise biological functions and how they contribute to the shared risk for psychiatric and IR-related conditions. Another limitation is the reliance on GWAS summary statistics derived from European ancestry populations, which may limit the generalisability of our findings to other populations. This issue highlights the need for more diverse genetic studies to ensure that our findings are applicable across different ethnic groups. The reliance on gene expression profiles from nervous tissues presents significant challenges, particularly given the often non-linear relationships between gene expression, protein function, and therapeutic efficacy (Munro et al., 2024). The T-SEM approach, while powerful in identifying tissue-specific gene expression effects across multiple genetically correlated traits, operates under the assumption that gene expression effects are consistent across all studied traits, potentially oversimplifying the complexity of biological interactions (Grotzinger et al., 2022a). In this respect, we employed the QGene statistic in an attempt to mitigate the risk of false-positive findings that could arise from such assumptions (Grotzinger et al., 2022a). However, the dynamic nature of gene regulation, epigenetic modifications, and the impact of environmental exposures can still exert tissue- or cell type-specific effects that might not be detected by our model (Pascual-Ahuir et al., 2020). Additionally, the drug repurposing results, while compelling, should be interpreted with much caution. Specific to drug repurposing, the relatively lower availability of human brain tissue samples remains a significant limitation. Moreover, the potential for off-target effects when repurposing drugs identified through gene expression overlaps must be carefully evaluated.

In conclusion, this study identified a common genetic factor underlying psychiatric and IR-related conditions, encapsulated by the Psych-IR multimorbidity factor. Overall, our findings suggest that the associated genetic factors are likely involved in pathways that regulate both brain function and metabolic processes, particularly during critical developmental windows. These findings have significant implications for our understanding of the co-occurrence between IR-related conditions and psychiatric disorders, providing new insights into the biological mechanisms that contribute to these comorbidities. Furthermore, the integration of genomic and transcriptomic data has identified potential candidate biomarkers and therapeutic targets, thereby providing the basis for the development of novel interventions. As research in this area continues to evolve, these findings have the potential to inform both scientific research and clinical practice, ultimately contributing to improved outcomes for patients with these co-occurring conditions.

## Data availability

Multivariate GWAS and TWAS summary statistics of the Psych-IR multimorbidity factor are available upon reasonable request to the authors.

## Ethical statement

We used publicly available summary statistics of genome-wide association studies conducted by external consortia, thus no specific authorization was required from the local Ethics Committee. No individual genotypic data were used or collected.

## Acknowledgments

This project has received funding from the European Union’s Horizon 2020 research and innovation programme under grant agreement No 847879 (PRIME, Prevention and Remediation of Insulin Multimorbidity in Europe). This publication is part of the projects ‘No labels needed: understanding psychiatric disorders based on genetic traits’ (with project number 09150161910091) and ’Advancing the discovery of genetic architecture across psychiatric disorders’ (with project number 09150162010073) of the research program Veni, which is partly financed by the Dutch Research Council (NWO) Health Research and Development (ZonMW). Research reported in this publication was supported by the National Institute Of Mental Health of the National Institutes of Health under Award Number R01MH124851. The content is solely the responsibility of the authors and does not necessarily represent the official views of any of the funders. This work was carried out on the Dutch national e-infrastructure with the support of SURF Cooperative. We also thank the researchers of the consortia that provided the univariate GWAS summary statistics used in our analyses and the participants of the cohorts to which they refer.

## Conflicts of interest

IHR is an employee and GP is the director and chief scientific officer of Drug Target ID, Ltd., but their activities at this company do not constitute competing interests with regard to this paper. AS has served as a consultant or speaker for Abbott, Abbvie, Angelini, AstraZeneca, Clinical Data, Boehringer, Bristol-Myers Squibb, Eli Lilly, GlaxoSmithKline, Innovapharma, Italfarmaco, Janssen, Lundbeck, Naurex, Pfizer, Polifarma, Sanofi, and Servier and Taliaz. During the past three years, JH and BF have received lecture honoraria as part of continuing medical education programs sponsored by Medice, all outside the submitted work. All the other authors declare no conflicts of interest.

## Notes

### Author Declarations

We used publicly available summary statistics of genome-wide association studies conducted by external consortia, thus no specific authorisation was required from the local Ethics Committee. No individual genotypic data were used or collected. Univariate GWAS summary statistics are available from: https://pgc.unc.edu/for-researchers/download-results/ https://www.ebi.ac.uk/gwas/publications/31589552 https://atlas.ctglab.nl/traitDB/3687 https://www.nature.com/articles/s41588-022-01058-3#MOESM1

