## Supplementary Tables S1-S6 and Supplementary Figures for "The multivariate genetic architecture of psychiatric and insulin resistance multimorbidity"

**Supplementary Information**

- Supplementary Table S3. Factor loadings for 2-factors model solution of the Exploratory Factor Analysis. 4
- Supplementary Table S4. Genetic correlations between the Psych-IR multimorbidity factor and 78 brain morphometric traits.......... 5
- Supplementary Table S5. Significant Independent Risk loci of Psych-IR multimorbidity factor (including significant QSNPs) and the univariate GWAS constituents.......... 8
- Supplementary Table S6. 150 independent risk loci for Psych-IR multimorbidity factor (after exclusion of significant Q-SNPs and those SNPs in LD with them) and their overlap in univariate GWAS constituents 12

### Supplementary Tables

**Supplementary Table S1. Exploratory Factor Analyses (based on odd autosomes)**

| Models | Variance explained | |
|---|---|---|
| | Total | Per factor |
| 1-factor | 0.314 | F1=0.314 |
| 2-factors | 0.521 | F1=0.321; F2=0.20 |
| 3-factors | 0.603 | F1=0.275; F2=0.176; F3=0.152 |

Note: Exploratory factor analyses were conducted on odd chromosomes using the *factanal* function of R with promax rotation. Models where each factor explained at least 20% of the variance were taken forward for confirmatory factor analyses.

**Supplementary Table S2. Model fit estimates of Confirmatory Factor Analyses**

| Models | Chromosomes | Chi2 | df | p_chisq | AIC | CFI | SRMR |
|---|---|---|---|---|---|---|---|
| 1-factor | Even autosomes | 514.419 | 20 | 2.78E-96 | 546.419 | 0.670 | 0.160 |
| 2-factors | Even autosomes | 56.562 | 15 | 9.73E-07 | 98.562 | 0.972 | 0.063 |
| **Final model** | | | | | | | |
| 2-factors | All autosomes | 78.559 | 15 | 1.28E-10 | 120.559 | 0.978 | 0.053 |

Note: Confirmatory Factor Analyses were conducted for 1-factor and 2-factor solutions using genetic correlations for even numbered chromosomes only and were based on the results of the exploratory factor analyses (which used odd chromosomes only). The 2-factors model solution was chosen as the best fit. The model fit of the chosen final model, estimated for all autosomes, is also shown above. df: degrees of freedom; AIC: Aikake Information Criterion; CFI: comparative fit index; SRMR: standardized root mean square residual.

**Supplementary Table S3. Factor loadings for 2-factors model solution of the Exploratory Factor Analysis**

| | Factor1 | Factor2 |
|---|---|---|
| ADHD | **0.476** | **0.483** |
| Anorexia | **-0.228** | **0.434** |
| OCD | **-0.342** | **0.501** |
| MDD | **0.28** | **0.841** |
| Schizophrenia | -0.108 | **0.464** |
| Metabolic Syndrome | **0.861** | <\|0.10\| |
| Obesity | **0.839** | <\|0.10\| |
| Type 2 diabetes | **0.796** | <\|0.10\| |

Note: Factor loadings were estimated using the odd autosomes genetic correlation matrix. This factor solution was the base for the confirmatory factor analysis final chosen model, where factors were assigned to traits when their standardized loading exceeded 0.20 (in bold).

**Supplementary Table S4. Genetic correlations between the Psych-IR multimorbidity factor and 78 brain morphometric traits**

| | rg | SE | P-value | $P\_Q_{TRAIT}$ |
|---|---|---|---|---|
| Subcortical volumes | | | | |
| accumbens | -0.068 | 0.038 | 0.0770 | 0.5880 |
| amygdala | 0.022 | 0.049 | 0.6542 | 0.0536 |
| brainstem | -0.080 | 0.033 | 0.0169 | 0.0146 |
| caudate | -0.029 | 0.031 | 0.3537 | 0.0145 |
| pallidum | -0.094 | 0.040 | 0.0175 | **1.71E-05** |
| putamen | 0.001 | 0.033 | 0.9666 | 0.4983 |
| thalamus | -0.056 | 0.038 | 0.1447 | 0.1115 |
| hippocampus | 0.015 | 0.048 | 0.7459 | 0.7105 |
| Cortical Surface Area (SA) | | | | |
| **SA_Total_SurfArea** | -0.151 | 0.033 | **4.89E-06** | 0.0009 |
| SA_bankssts | -0.138 | 0.044 | 0.0015 | 0.0467 |
| SA_caudalanteriorcingulate | -0.044 | 0.039 | 0.2565 | 0.5934 |
| SA_caudalmiddlefrontal | 0.014 | 0.035 | 0.6865 | 0.0387 |
| SA_cuneus | 0.024 | 0.038 | 0.5189 | 0.6560 |
| SA_entorhinal | -0.027 | 0.043 | 0.5321 | 0.3527 |
| SA_frontalpole | 0.021 | 0.047 | 0.6598 | **5.83E-07** |
| SA_fusiform | -0.053 | 0.041 | 0.1913 | 0.0902 |
| SA_inferiorparietal | -0.109 | 0.041 | 0.0082 | 0.4540 |
| **SA_inferiortemporal** | -0.183 | 0.045 | **4.60E-05** | 0.2337 |
| SA_insula | 0.023 | 0.031 | 0.4656 | 0.4764 |
| SA_isthmuscingulate | 0.153 | 0.046 | 7.88E-04 | 0.7201 |
| **SA_lateraloccipital** | 0.113 | 0.032 | **0.0005** | 0.3515 |
| SA_lateralorbitofrontal | 0.065 | 0.036 | 0.0691 | 0.0543 |
| SA_lingual | 0.038 | 0.033 | 0.2492 | 0.2089 |
| SA_medialorbitofrontal | -0.003 | 0.048 | 0.9577 | **9.71E-07** |
| SA_middletemporal | -0.107 | 0.036 | 0.0027 | 0.2144 |
| SA_paracentral | 0.000 | 0.042 | 0.9993 | 0.9342 |
| SA_parahippocampal | -0.083 | 0.040 | 0.0387 | **5.76E-08** |
| SA_parsopercularis | 0.060 | 0.037 | 0.1078 | 0.7616 |
| SA_parsorbitalis | -0.027 | 0.035 | 0.4482 | 0.1239 |
| SA_parstriangularis | 0.024 | 0.033 | 0.4678 | 0.7031 |
| SA_pericalcarine | 0.045 | 0.031 | 0.1425 | 0.0054 |
| SA_postcentral | -0.103 | 0.039 | 0.0082 | 0.0018 |
| SA_posteriorcingulate | -0.039 | 0.041 | 0.3392 | 0.9951 |
| SA_precentral | -0.048 | 0.040 | 0.2279 | 0.0447 |
| SA_precuneus | 0.062 | 0.037 | 0.0924 | 0.2789 |
| SA_rostralanteriorcingulate | 0.004 | 0.041 | 0.9168 | 0.3238 |
| SA_rostralmiddlefrontal | 0.003 | 0.035 | 0.9410 | 0.2969 |
| SA_superiorfrontal | 0.026 | 0.038 | 0.4843 | 0.9356 |
| SA_superiorparietal | 0.056 | 0.040 | 0.1590 | 0.2318 |

| | | | | |
|---|---|---|---|---|
| SA_superiortemporal | -0.048 | 0.033 | 0.1399 | 0.2666 |
| SA_supramarginal | -0.034 | 0.038 | 0.3681 | 0.8739 |
| SA_temporalpole | 0.062 | 0.039 | 0.1130 | 1.0000 |
| SA_transversetemporal | 0.037 | 0.036 | 0.3123 | 0.0889 |
| Cortical Thickness (CT) | | | | |
| CT_Total_Thickness | -0.093 | 0.033 | 0.0052 | 0.8980 |
| CT_bankssts | -0.101 | 0.059 | 0.0880 | **1.01E-07** |
| CT_caudalanteriorcingulate | -0.002 | 0.041 | 0.9684 | 0.9828 |
| CT_caudalmiddlefrontal | 0.076 | 0.042 | 0.0728 | **0.0002** |
| CT_cuneus | 0.074 | 0.046 | 0.1063 | 0.0678 |
| CT_entorhinal | -0.084 | 0.043 | 0.0527 | **1.19E-10** |
| CT_frontalpole | -0.077 | 0.060 | 0.1967 | **1.46E-10** |
| CT_fusiform | -0.135 | 0.060 | 0.0245 | **5.01E-08** |
| CT_inferiorparietal | 0.117 | 0.053 | 0.0278 | 0.2023 |
| CT_inferiortemporal | -0.080 | 0.044 | 0.0714 | 0.8568 |
| CT_insula | -0.029 | 0.036 | 0.4201 | 0.2928 |
| CT_isthmuscingulate | 0.019 | 0.032 | 0.5547 | 0.0253 |
| CT_lateraloccipital | 0.037 | 0.047 | 0.4337 | 0.3950 |
| CT_lateralorbitofrontal | -0.029 | 0.044 | 0.5082 | 0.5629 |
| CT_lingual | 0.027 | 0.034 | 0.4240 | 0.1232 |
| CT_medialorbitofrontal | 0.032 | 0.039 | 0.4089 | **0.0002** |
| CT_middletemporal | -0.079 | 0.041 | 0.0536 | 0.3095 |
| CT_paracentral | 0.008 | 0.046 | 0.8632 | 0.0590 |
| CT_parahippocampal | -0.008 | 0.040 | 0.8365 | **1.46E-05** |
| CT_parsopercularis | 0.023 | 0.060 | 0.7078 | 0.0104 |
| CT_parsorbitalis | 0.021 | 0.054 | 0.7051 | **3.59E-07** |
| CT_parstriangularis | 0.069 | 0.046 | 0.1356 | 0.0129 |
| CT_pericalcarine | 0.124 | 0.049 | 0.0113 | 0.7679 |
| CT_postcentral | 0.054 | 0.037 | 0.1466 | 0.9558 |
| CT_posteriorcingulate | -0.006 | 0.038 | 0.8689 | 0.0107 |
| CT_precentral | -0.039 | 0.047 | 0.4065 | 0.0114 |
| CT_precuneus | 0.034 | 0.043 | 0.4382 | 0.4083 |
| CT_rostralanteriorcingulate | 0.121 | 0.045 | 0.0071 | 0.2303 |
| CT_rostralmiddlefrontal | -0.017 | 0.049 | 0.7311 | 0.0966 |
| CT_superiorfrontal | 0.029 | 0.049 | 0.5563 | 0.3101 |
| CT_superiorparietal | 0.106 | 0.042 | 0.0119 | 0.1143 |
| CT_superiortemporal | -0.047 | 0.037 | 0.2010 | 0.9705 |
| CT_supramarginal | 0.070 | 0.041 | 0.0856 | 0.5351 |
| CT_temporalpole | -0.135 | 0.071 | 0.0579 | **1.42E-08** |
| CT_transversetemporal | 0.022 | 0.040 | 0.5751 | 0.4864 |

Note: Genetic correlations (rg) with the Psych-IR factor were estimated using Genomic SEM. $P\_Q_{trait}$ column indexes whether there is significant heterogeneity with respect to the genetic correlations at the level of the individual disorders/conditions (i.e. factor indicators). Significant associations of brain traits and significant $Q_{trait}$ heterogeneity index are shown in bold. Bonferroni significance threshold is $P_{Bonf}$=6.41E-04 (0.05/78 brain traits).

**Supplementary Table S5. Significant Independent Risk loci of Psych-IR multimorbidity factor (including significant $Q_{SNPs}$) and the univariate GWAS constituents**

| Locus | Locus Unique ID | Index SNP | CHR | BP | Start | End | P-value | N Ind. Sig. SNPs | $Q_{SNP}$_hit |
|---|---|---|---|---|---|---|---|---|---|
| 1 | 1:26987646:A:G | rs193084249 | 1 | 26987646 | 26933591 | 27335529 | 1.94E-09 | 2 | - |
| 2 | 1:40035928:G:T | rs3768321 | 1 | 40035928 | 39551488 | 40161734 | 7.59E-22 | 14 | - |
| 3 | 1:62579891:G:T | rs12140153 | 1 | 62579891 | 62579891 | 62579891 | 1.86E-11 | 1 | - |
| 4 | 1:72837490:C:T | rs11209951 | 1 | 72837490 | 72748669 | 72940273 | 4.01E-08 | 1 | - |
| 5 | 1:177913519:C:T | rs10913469 | 1 | 177913519 | 177792715 | 177936599 | 7.34E-15 | 9 | - |
| 6 | 1:201884952:C:T | rs2819348 | 1 | 201884952 | 201787940 | 201887457 | 2.13E-08 | 1 | - |
| 7 | 1:210306846:A:G | rs4844949 | 1 | 210306846 | 210068954 | 210306846 | 3.95E-09 | 1 | - |
| 8 | 1:214159256:C:T | rs340874 | 1 | 214159256 | 213995780 | 214192133 | 5.01E-15 | 6 | - |
| 9 | 1:219748818:C:G | rs2820446 | 1 | 219748818 | 219622596 | 219798632 | 1.80E-11 | 5 | - |
| 10 | 1:230297778:A:T | rs2281718 | 1 | 230297778 | 230278291 | 230326412 | 1.44E-14 | 4 | - |
| 11 | 2:633659:C:T | rs13035713 | 2 | 633659 | 408713 | 699392 | 1.77E-14 | 4 | - |
| 12 | 2:21221035:A:C | rs4665710 | 2 | 21221035 | 21121464 | 21245329 | 1.03E-10 | 1 | $Q_{SNP}$_hit |
| 13 | 2:43469615:A:G | rs80323638 | 2 | 43469615 | 43449385 | 43864089 | 2.51E-10 | 2 | $Q_{SNP}$_hit |
| 14 | 2:58933591:C:T | rs1861410 | 2 | 58933591 | 58760169 | 59337008 | 1.25E-12 | 11 | - |
| 15 | 2:60585806:C:T | rs243019 | 2 | 60585806 | 60565568 | 60586707 | 3.07E-09 | 1 | - |
| 16 | 2:65276049:C:T | rs1009360 | 2 | 65276049 | 65235333 | 65705581 | 2.38E-13 | 5 | - |
| 17 | 2:86807854:A:C | rs62147190 | 2 | 86807854 | 86648492 | 86826508 | 8.07E-09 | 1 | - |
| 18 | 2:100729293:C:T | rs4851250 | 2 | 100729293 | 100576304 | 100871361 | 3.73E-10 | 4 | - |
| 19 | 2:161087411:C:T | rs10181181 | 2 | 161087411 | 161087411 | 161277857 | 6.83E-10 | 2 | - |
| 20 | 2:165532454:A:G | rs10187501 | 2 | 165532454 | 165501849 | 165731664 | 2.07E-13 | 5 | - |
| 21 | 2:171631258:G:T | rs4668314 | 2 | 171631258 | 171570488 | 171680587 | 1.37E-08 | 1 | - |
| 22 | 2:175241566:A:G | rs72917544 | 2 | 175241566 | 175236016 | 175241566 | 8.79E-10 | 1 | - |
| 23 | 2:181607676:A:C | rs9630985 | 2 | 181607676 | 181518061 | 181618654 | 4.14E-10 | 2 | - |
| 24 | 2:227101411:A:G | rs2972144 | 2 | 227101411 | 226813090 | 227199263 | 2.48E-24 | 17 | - |
| 25 | 2:228998742:A:G | rs72967047 | 2 | 228998742 | 228971784 | 229021719 | 5.55E-12 | 3 | - |
| 26 | 2:230814719:C:T | rs6722477 | 2 | 230814719 | 230616197 | 230854290 | 4.56E-08 | 1 | - |
| 27 | 3:12329783:C:T | rs17036160 | 3 | 12329783 | 12108754 | 12413339 | 3.22E-10 | 2 | - |
| 28 | 3:15718652:A:T | rs2470540 | 3 | 15718652 | 15680525 | 15970334 | 7.74E-10 | 2 | - |
| 29 | 3:50041313:C:T | rs6765484 | 3 | 50041313 | 49669948 | 50248954 | 1.11E-15 | 9 | - |
| 30 | 3:53125585:C:T | rs2564921 | 3 | 53125585 | 52970877 | 53139977 | 9.50E-09 | 1 | - |
| 31 | 3:62459819:A:C | rs76824303 | 3 | 62459819 | 62459819 | 62459819 | 3.43E-08 | 1 | - |
| 32 | 3:63897215:C:T | rs2292662 | 3 | 63897215 | 63853423 | 64007214 | 3.63E-11 | 2 | - |
| 33 | 3:94038085:C:G | rs1454687 | 3 | 94038085 | 93883640 | 94198182 | 1.91E-10 | 2 | - |
| 34 | 3:123065778:A:G | rs11708067 | 3 | 123065778 | 123051019 | 123131254 | 2.15E-09 | 1 | $Q_{SNP}$_hit |
| 35 | 3:131632210:C:T | rs1228588 | 3 | 131632210 | 131447660 | 131792411 | 6.00E-11 | 2 | - |
| 36 | 3:135955604:C:T | rs895893 | 3 | 135955604 | 135656190 | 136752590 | 2.54E-17 | 11 | - |
| 37 | 3:150097635:C:G | rs9844972 | 3 | 150097635 | 150066540 | 150097635 | 2.65E-09 | 1 | - |
| 38 | 3:173117548:G:T | rs546738 | 3 | 173117548 | 173044559 | 173127808 | 4.18E-09 | 1 | - |
| 39 | 3:185520948:A:G | rs9854769 | 3 | 185520948 | 185459675 | 185538006 | 4.67E-18 | 3 | $Q_{SNP}$_hit |
| 40 | 4:3241845:C:T | rs362307 | 4 | 3241845 | 3142660 | 3422211 | 1.17E-09 | 4 | - |
| 41 | 4:6304286:C:T | rs1046319 | 4 | 6304286 | 6263996 | 6328507 | 5.78E-15 | 7 | $Q_{SNP}$_hit |
| 42 | 4:45175691:C:T | rs13130484 | 4 | 45175691 | 45068929 | 45187622 | 3.26E-11 | 2 | - |
| 43 | 4:67802428:C:T | rs12509944 | 4 | 67802428 | 67802098 | 67903330 | 5.12E-09 | 1 | - |
| 44 | 4:96170439:C:T | rs4699837 | 4 | 96170439 | 96123023 | 96181240 | 4.36E-08 | 1 | - |

| | | | | | | | | | |
|---|---|---|---|---|---|---|---|---|---|
| 45 | 4:103679117:G:T | rs223490 | 4 | 103679117 | 103557311 | 104378012 | 1.37E-10 | 8 | - |
| 46 | 4:137094048:A:C | rs1724557 | 4 | 137094048 | 136966779 | 137098470 | 9.24E-11 | 1 | - |
| 47 | 4:140892872:C:T | rs1397440 | 4 | 140892872 | 140858717 | 140948095 | 4.02E-09 | 1 | - |
| 48 | 5:53271420:A:G | rs702634 | 5 | 53271420 | 53271420 | 53287099 | 1.53E-09 | 1 | - |
| 49 | 5:55861894:A:G | rs9687846 | 5 | 55861894 | 55794632 | 55876283 | 6.01E-16 | 5 | - |
| 50 | 5:57607142:A:T | rs151912 | 5 | 57607142 | 57527886 | 57636145 | 3.23E-08 | 1 | - |
| 51 | 5:75003678:C:T | rs2307111 | 5 | 75003678 | 74568235 | 75038901 | 2.48E-15 | 6 | - |
| 52 | 5:78522895:C:T | rs10078815 | 5 | 78522895 | 78416416 | 78628979 | 1.40E-10 | 1 | - |
| 53 | 5:87703099:C:G | rs7445169 | 5 | 87703099 | 87440497 | 88010829 | 7.28E-11 | 2 | - |
| 54 | 5:102422968:C:T | rs115505614 | 5 | 102422968 | 101209723 | 102726073 | 1.27E-10 | 2 | $Q_{SNP}$_hit |
| 55 | 5:103995368:A:G | rs325485 | 5 | 103995368 | 103791044 | 104055261 | 2.88E-08 | 1 | - |
| 56 | 5:118715275:G:T | rs924623 | 5 | 118715275 | 118697849 | 118800074 | 2.09E-08 | 1 | - |
| 57 | 6:7275941:C:T | rs11243150 | 6 | 7275941 | 7240876 | 7306153 | 5.12E-09 | 2 | - |
| 58 | 6:20673880:C:T | rs7451008 | 6 | 20673880 | 20483407 | 20896553 | 6.02E-22 | 11 | - |
| 59 | 6:26200677:A:G | rs806794 | 6 | 26200677 | 26180634 | 26259807 | 3.19E-09 | 1 | - |
| 60 | 6:33546498:C:T | rs5745582 | 6 | 33546498 | 33463961 | 33803752 | 1.77E-09 | 6 | - |
| 61 | 6:34749849:A:G | rs9689160 | 6 | 34749849 | 34548206 | 35162141 | 1.10E-08 | 2 | - |
| 62 | 6:40369081:C:T | rs34045288 | 6 | 40369081 | 40348653 | 40383533 | 3.31E-08 | 1 | - |
| 63 | 6:43757896:A:C | rs998584 | 6 | 43757896 | 43756863 | 43765533 | 2.01E-16 | 2 | - |
| 64 | 6:50788778:A:C | rs3798519 | 6 | 50788778 | 50664180 | 50940677 | 3.41E-13 | 4 | - |
| 65 | 6:79349892:A:C | rs236852 | 6 | 79349892 | 79316046 | 79477484 | 4.24E-08 | 1 | - |
| 66 | 6:127454893:A:G | rs72959041 | 6 | 127454893 | 127439897 | 127529780 | 9.91E-09 | 1 | - |
| 67 | 6:139835329:A:T | rs2982521 | 6 | 139835329 | 139828916 | 139844429 | 1.34E-09 | 1 | - |
| 68 | 6:153371503:C:T | rs7761831 | 6 | 153371503 | 153350709 | 153449670 | 2.36E-10 | 3 | - |
| 69 | 7:1920356:A:G | rs34040190 | 7 | 1920356 | 1824028 | 2110850 | 8.59E-11 | 5 | - |
| 70 | 7:2760750:A:C | rs798549 | 7 | 2760750 | 2760750 | 2862891 | 2.72E-08 | 1 | - |
| 71 | 7:15040241:C:T | rs2189723 | 7 | 15040241 | 15024954 | 15065983 | 2.19E-08 | 1 | $Q_{SNP}$_hit |
| 72 | 7:17287998:A:G | rs2106727 | 7 | 17287998 | 17281007 | 17309279 | 9.10E-09 | 1 | - |
| 73 | 7:28198677:C:T | rs1708302 | 7 | 28198677 | 28142088 | 28256240 | 2.84E-16 | 6 | - |
| 74 | 7:44255643:A:G | rs878521 | 7 | 44255643 | 44223721 | 44265179 | 5.47E-10 | 1 | - |
| 75 | 7:50683678:A:G | rs2715126 | 7 | 50683678 | 50659193 | 50685625 | 5.91E-09 | 1 | - |
| 76 | 7:130466091:A:G | rs1447060 | 7 | 130466091 | 130422934 | 130468190 | 3.89E-13 | 3 | - |
| 77 | 7:150657095:A:G | rs56282717 | 7 | 150657095 | 150610935 | 150660239 | 8.23E-10 | 1 | - |
| 78 | 8:9183358:A:G | rs9987289 | 8 | 9183358 | 9177732 | 9231661 | 1.38E-11 | 2 | - |
| 79 | 8:9861116:C:T | rs6601406 | 8 | 9861116 | 9860588 | 10005526 | 8.21E-10 | 2 | - |
| 80 | 8:10675500:C:G | rs2409656 | 8 | 10675500 | 10570877 | 10708509 | 5.19E-09 | 1 | - |
| 81 | 8:19803093:C:T | rs3779788 | 8 | 19803093 | 19526176 | 19974789 | 4.72E-27 | 59 | - |
| 82 | 8:30863938:C:T | rs10954772 | 8 | 30863938 | 30819854 | 30879207 | 3.12E-12 | 3 | - |
| 83 | 8:41519462:A:G | rs515071 | 8 | 41519462 | 41488038 | 41537318 | 1.68E-08 | 1 | $Q_{SNP}$_hit |
| 84 | 8:116560619:C:T | rs7825679 | 8 | 116560619 | 116409058 | 116645056 | 9.66E-13 | 3 | - |
| 85 | 8:118185025:A:G | rs3802177 | 8 | 118185025 | 118184783 | 118220270 | 2.62E-17 | 2 | $Q_{SNP}$_hit |
| 86 | 8:126480367:C:T | rs4871603 | 8 | 126480367 | 126474306 | 126648243 | 6.52E-15 | 4 | - |
| 87 | 9:22134068:A:G | rs10811660 | 9 | 22134068 | 22129579 | 22137685 | 3.34E-20 | 3 | $Q_{SNP}$_hit |
| 88 | 9:28410683:C:T | rs1412234 | 9 | 28410683 | 28410683 | 28488365 | 2.51E-09 | 1 | - |
| 89 | 9:134849037:A:G | rs11243586 | 9 | 134849037 | 134802170 | 134946805 | 1.00E-08 | 1 | - |
| 90 | 10:12321633:C:T | rs7394200 | 10 | 12321633 | 12242326 | 12328010 | 5.64E-12 | 5 | - |
| 91 | 10:64988931:A:T | rs7916868 | 10 | 64988931 | 64874754 | 65400352 | 2.63E-12 | 5 | - |
| 92 | 10:80948593:A:G | rs703974 | 10 | 80948593 | 80906729 | 81067040 | 1.08E-15 | 11 | - |
| 93 | 10:94465559:C:T | rs5015480 | 10 | 94465559 | 93544818 | 94500111 | 1.30E-21 | 18 | - |
| 94 | 10:99772404:A:G | rs563296 | 10 | 99772404 | 99762693 | 99784552 | 5.25E-09 | 1 | - |

| | | | | | | | | | |
|---|---|---|---|---|---|---|---|---|---|
| 95 | 10:113940329:C:T | rs2792751 | 10 | 113940329 | 113901194 | 113950418 | 1.65E-08 | 1 | - |
| 96 | 10:114748617:A:G | rs4074718 | 10 | 114748617 | 114722134 | 114867427 | 4.74E-29 | 14 | $Q_{SNP}$_hit |
| 97 | 11:2857194:A:C | rs2237895 | 11 | 2857194 | 2836213 | 2857897 | 3.08E-14 | 4 | - |
| 98 | 11:8694830:A:G | rs10128597 | 11 | 8694830 | 8440969 | 8694830 | 2.56E-09 | 1 | - |
| 99 | 11:13269921:A:T | rs11825504 | 11 | 13269921 | 13268386 | 13350131 | 1.56E-08 | 2 | - |
| 100 | 11:17415190:C:G | rs4148646 | 11 | 17415190 | 17368013 | 17421886 | 1.97E-09 | 1 | $Q_{SNP}$_hit |
| 101 | 11:27722298:G:T | rs7944119 | 11 | 27722298 | 27477864 | 27748493 | 3.65E-12 | 4 | - |
| 102 | 11:43752522:A:G | rs10838158 | 11 | 43752522 | 43634973 | 43878534 | 9.39E-09 | 1 | - |
| 103 | 11:47529947:A:C | rs7124681 | 11 | 47529947 | 47232038 | 47946836 | 9.43E-16 | 4 | - |
| 104 | 11:57268252:C:T | rs112450479 | 11 | 57268252 | 57158320 | 57386178 | 4.73E-08 | 1 | - |
| 105 | 11:65662752:A:G | rs588114 | 11 | 65662752 | 65211979 | 65663547 | 2.63E-10 | 4 | - |
| 106 | 11:72460694:C:T | rs11603349 | 11 | 72460694 | 72419514 | 72851463 | 5.71E-09 | 1 | $Q_{SNP}$_hit |
| 107 | 11:76485358:A:G | rs737185 | 11 | 76485358 | 76464812 | 76511271 | 7.51E-09 | 1 | - |
| 108 | 11:92708710:C:G | rs10830963 | 11 | 92708710 | 92667730 | 92727089 | 8.89E-17 | 4 | $Q_{SNP}$_hit |
| 109 | 11:102948592:A:G | rs2510087 | 11 | 102948592 | 102929625 | 103153310 | 2.64E-08 | 1 | - |
| 110 | 11:116702123:C:T | rs5141 | 11 | 116702123 | 116519358 | 117175658 | 4.77E-21 | 29 | - |
| 111 | 11:118949083:A:G | rs1177563 | 11 | 118949083 | 118913993 | 118958869 | 5.58E-10 | 1 | - |
| 112 | 12:26320656:C:T | rs10842685 | 12 | 26320656 | 26313849 | 26339986 | 2.70E-08 | 1 | - |
| 113 | 12:27919145:C:T | rs9668691 | 12 | 27919145 | 27905969 | 27965150 | 9.80E-09 | 1 | - |
| 114 | 12:50263148:A:G | rs7132908 | 12 | 50263148 | 50240304 | 50285780 | 1.62E-10 | 2 | - |
| 115 | 12:56467865:C:G | rs10876869 | 12 | 56467865 | 56368078 | 56609885 | 9.06E-09 | 1 | - |
| 116 | 12:66221060:A:T | rs2258238 | 12 | 66221060 | 66165471 | 66281348 | 4.48E-09 | 1 | $Q_{SNP}$_hit |
| 117 | 12:108618630:C:T | rs3764002 | 12 | 108618630 | 108609634 | 108629780 | 1.65E-10 | 1 | - |
| 118 | 12:121451910:C:T | rs11065397 | 12 | 121451910 | 121432117 | 121463562 | 7.41E-09 | 1 | $Q_{SNP}$_hit |
| 119 | 12:123676763:A:G | rs2102949 | 12 | 123676763 | 123296204 | 123913433 | 6.38E-11 | 6 | - |
| 120 | 12:124409502:A:G | rs7133378 | 12 | 124409502 | 124401710 | 124496316 | 5.27E-09 | 1 | - |
| 121 | 13:58628636:C:T | rs8000972 | 13 | 58628636 | 58254773 | 58828747 | 2.64E-10 | 2 | - |
| 122 | 13:80709595:G:T | rs12428731 | 13 | 80709595 | 80620642 | 80758943 | 1.51E-12 | 3 | - |
| 123 | 14:23823913:A:T | rs7148564 | 14 | 23823913 | 23792416 | 23837484 | 3.26E-08 | 1 | - |
| 124 | 14:25930988:A:C | rs8015400 | 14 | 25930988 | 25927298 | 25994917 | 2.99E-08 | 1 | - |
| 125 | 14:79891882:A:G | rs10150482 | 14 | 79891882 | 79833494 | 79945162 | 4.58E-10 | 1 | - |
| 126 | 15:41855736:G:T | rs1023193 | 15 | 41855736 | 41798762 | 42190692 | 1.90E-11 | 7 | - |
| 127 | 15:51893161:C:T | rs4774595 | 15 | 51893161 | 51709536 | 52004250 | 3.78E-09 | 1 | - |
| 128 | 15:56870157:C:T | rs139280016 | 15 | 56870157 | 56825676 | 57633098 | 4.93E-10 | 3 | - |
| 129 | 15:58678720:C:T | rs261290 | 15 | 58678720 | 58671559 | 58741891 | 3.37E-16 | 9 | - |
| 130 | 15:63922474:C:T | rs11071759 | 15 | 63922474 | 63798569 | 64136472 | 1.05E-09 | 2 | - |
| 131 | 15:68080886:A:T | rs4776970 | 15 | 68080886 | 67848128 | 68113771 | 1.48E-09 | 1 | - |
| 132 | 15:73451443:A:C | rs1904335 | 15 | 73451443 | 73392760 | 73621435 | 1.34E-09 | 1 | - |
| 133 | 15:77837055:G:T | rs62009090 | 15 | 77837055 | 77310345 | 77912717 | 8.23E-17 | 10 | - |
| 134 | 15:91511260:A:G | rs12910825 | 15 | 91511260 | 91502524 | 91537151 | 2.09E-08 | 1 | $Q_{SNP}$_hit |
| 135 | 16:387037:A:C | rs7498932 | 16 | 387037 | 376836 | 412974 | 2.29E-08 | 1 | - |
| 136 | 16:24735736:C:T | rs200538 | 16 | 24735736 | 24709089 | 24832693 | 2.05E-08 | 2 | - |
| 137 | 16:28917644:C:T | rs7188071 | 16 | 28917644 | 28403471 | 28955702 | 3.29E-09 | 1 | - |
| 138 | 16:30018720:C:T | rs12921753 | 16 | 30018720 | 29923510 | 30147265 | 4.32E-15 | 4 | - |
| 139 | 16:31131174:A:G | rs4889620 | 16 | 31131174 | 30916233 | 31149142 | 1.53E-11 | 1 | - |

| | | | | | | | | | |
|---|---|---|---|---|---|---|---|---|---|
| 140 | 16:53806453:A:G | rs56094641 | 16 | 53806453 | 53797908 | 53848561 | 3.00E-36 | 5 | - |
| 141 | 16:57017319:A:G | rs1800777 | 16 | 57017319 | 56742475 | 57049137 | 3.88E-18 | 13 | - |
| 142 | 16:69651866:C:T | rs862320 | 16 | 69651866 | 69547741 | 70112729 | 1.75E-14 | 7 | - |
| 143 | 16:81534790:C:T | rs2925979 | 16 | 81534790 | 81449060 | 81549708 | 1.18E-15 | 7 | - |
| 144 | 17:1830836:C:T | rs77483079 | 17 | 1830836 | 1820080 | 1859542 | 1.25E-09 | 2 | - |
| 145 | 17:3882309:A:G | rs17763551 | 17 | 3882309 | 3880546 | 4165741 | 3.58E-11 | 3 | - |
| 146 | 17:29720140:C:T | rs8065496 | 17 | 29720140 | 29470288 | 29735829 | 6.15E-09 | 1 | - |
| 147 | 17:34850623:G:T | rs7222903 | 17 | 34850623 | 34825861 | 34961772 | 5.00E-09 | 2 | - |
| 148 | 17:36103565:A:G | rs11263763 | 17 | 36103565 | 36096300 | 36104121 | 3.50E-08 | 1 | $Q_{SNP}$_hit |
| 149 | 17:40721042:C:T | rs650558 | 17 | 40721042 | 40565200 | 41018481 | 6.22E-12 | 2 | - |
| 150 | 17:41926126:C:T | rs72836561 | 17 | 41926126 | 41809207 | 42282225 | 4.53E-16 | 3 | - |
| 151 | 17:46124326:C:G | rs9900074 | 17 | 46124326 | 45986496 | 46323934 | 7.71E-09 | 1 | - |
| 152 | 17:47090785:C:T | rs11079849 | 17 | 47090785 | 46835629 | 47145848 | 2.40E-11 | 6 | - |
| 153 | 17:65832576:A:G | rs12452511 | 17 | 65832576 | 65764164 | 66096529 | 9.23E-13 | 4 | - |
| 154 | 17:76405736:C:T | rs4969143 | 17 | 76405736 | 76386037 | 76406170 | 1.57E-08 | 1 | - |
| 155 | 17:76798155:C:T | rs2306527 | 17 | 76798155 | 76661269 | 76833916 | 2.69E-08 | 1 | - |
| 156 | 18:1839911:G:T | rs60764613 | 18 | 1839911 | 1811604 | 1914051 | 2.76E-08 | 1 | - |
| 157 | 18:21134239:C:T | rs1631685 | 18 | 21134239 | 21074255 | 21165409 | 7.39E-11 | 2 | - |
| 158 | 18:57848651:A:G | rs66922415 | 18 | 57848651 | 57728947 | 58090217 | 1.01E-20 | 15 | - |
| 159 | 19:7970635:A:G | rs4804833 | 19 | 7970635 | 7931242 | 7978430 | 3.59E-11 | 3 | - |
| 160 | 19:8429323:A:G | rs116843064 | 19 | 8429323 | 8429323 | 8441959 | 2.50E-18 | 2 | - |
| 161 | 19:33890838:C:G | rs10406327 | 19 | 33890838 | 33890838 | 33897478 | 1.97E-08 | 1 | - |
| 162 | 19:46157237:A:G | rs10407429 | 19 | 46157237 | 46108455 | 46201213 | 4.33E-13 | 5 | - |
| 163 | 19:47569003:A:G | rs3810291 | 19 | 47569003 | 47559074 | 47623080 | 1.71E-14 | 1 | - |
| 164 | 20:32657378:C:T | rs932388 | 20 | 32657378 | 32514061 | 32729444 | 1.90E-08 | 1 | - |
| 165 | 20:43042364:C:T | rs1800961 | 20 | 43042364 | 42958768 | 43042364 | 1.32E-16 | 2 | - |
| 166 | 20:50986299:A:G | rs13037010 | 20 | 50986299 | 50809213 | 51264331 | 1.84E-12 | 8 | - |
| 167 | 20:62450664:C:T | rs6011155 | 20 | 62450664 | 62402860 | 62471527 | 2.21E-08 | 1 | - |
| 168 | 21:46494995:C:T | rs9977825 | 21 | 46494995 | 46487831 | 46558780 | 4.32E-08 | 1 | - |

Note. Genomic risk loci were identified using FUMA (v1.5.6; Watanabe et al 2017). BP refers to the base pair position of the index SNP. Start and End columns refer to the BP positions of the borders of the associated genomic loci. $Q_{SNP}$_hit column indicates loci that overlap with genome-wide significant $Q_{SNPs}$ loci for the Psych-IR factor and that are no longer present after the exclusion of significant $Q_{SNPs}$ (i.e., in **Supplementary Table S6**). N Indep. Sig. SNPs=Number of genome-wide significant independent (at r2 < 0.6) SNPs within each locus.

**Supplementary Table S6. 150 independent risk loci for Psych-IR multimorbidity factor (after exclusion of significant Q-SNPs and those SNPs in LD with them) and their overlap in univariate GWAS constituents**

| Locus | Locus Unique ID | Index SNP | CHR | BP | Start | End | P-value | Overlap with Univariate GWAS Loci | Independent Significant SNPs |
|---|---|---|---|---|---|---|---|---|---|
| 1 | 1:26987646:A:G | rs193084249 | 1 | 26987646 | 26933591 | 27335529 | 1.94E-09 | MetS | rs193084249;rs182050989 |
| 2 | 1:40035928:G:T | rs3768321 | 1 | 40035928 | 39551488 | 40161734 | 7.59E-22 | MetS; T2DM | rs3768321;rs77027633;rs2310799;rs3916164;rs116401702;rs147860177;rs13374459;rs6684217;rs111623477;rs72661980;rs61779313;rs12402990;rs1180333;rs72665235 |
| 3 | 1:62579891:G:T | rs12140153 | 1 | 62579891 | 62579891 | 62579891 | 1.86E-11 | T2DM | rs12140153 |
| 4 | 1:72837490:C:T | rs11209951 | 1 | 72837490 | 72748669 | 72940273 | 4.01E-08 | MDD | rs11209951 |
| 5 | 1:177913519:C:T | rs10913469 | 1 | 177913519 | 177792715 | 177936599 | 7.34E-15 | MetS; T2DM | rs10913469;rs1336783;rs10798585;rs12747656;rs12757154;rs34472009;rs1336780;rs575908;rs591120 |
| 6 | 1:201884952:C:T | rs2819348 | 1 | 201884952 | 201787940 | 201887457 | 2.13E-08 | - | rs2819348 |
| 7 | 1:210306846:A:G | rs4844949 | 1 | 210306846 | 210068954 | 210306846 | 3.95E-09 | - | rs4844949 |
| 8 | 1:214145731:C:G | rs340882 | 1 | 214145731 | 213995780 | 214192133 | 9.34E-15 | T2DM | rs6540803;rs340882;rs340839;rs3767845;rs72753599;rs340868 |
| 9 | 1:219748818:C:G | rs2820446 | 1 | 219748818 | 219622596 | 219798632 | 1.80E-11 | T2DM | rs2820446;rs12133396;rs2066152;rs4323719;rs10779360 |
| 10 | 1:230302521:G:T | rs4846840 | 1 | 230302521 | 230278291 | 230326412 | 9.77E-12 | MetS | rs4846840;rs612577;rs1555289 |
| 11 | 2:633659:C:T | rs13035713 | 2 | 633659 | 408713 | 699392 | 1.77E-14 | MetS; T2DM | rs62107261;rs13035713;rs2867121;rs12999373 |
| 12 | 2:58933591:C:T | rs1861410 | 2 | 58933591 | 58760169 | 59337008 | 1.25E-12 | MetS; T2DM | rs1861410;rs2708149;rs1641155;rs10210385;rs6719884;rs764975;rs7584990;rs7572387;rs12472381;rs6753852;rs66517511 |
| 13 | 2:60580831:A:G | rs9309325 | 2 | 60580831 | 60578843 | 60585806 | 8.14E-09 | T2DM | rs9309325 |
| 14 | 2:65276049:C:T | rs1009360 | 2 | 65276049 | 65235333 | 65386462 | 2.38E-13 | MetS; T2DM | rs1009360;rs71424153;rs12472718;rs2540953 |
| 15 | 2:86807854:A:C | rs62147190 | 2 | 86807854 | 86648492 | 86826508 | 8.07E-09 | - | rs62147190 |
| 16 | 2:100729293:C:T | rs4851250 | 2 | 100729293 | 100576304 | 100871361 | 3.73E-10 | - | rs4851250;rs13002946;rs7566527;rs56909870 |
| 17 | 2:161087411:C:T | rs10181181 | 2 | 161087411 | 161087411 | 161277857 | 6.83E-10 | T2DM | rs10181181;rs12692596 |
| 18 | 2:165532454:A:G | rs10187501 | 2 | 165532454 | 165501849 | 165731664 | 2.07E-13 | MetS; T2DM | rs10187501;rs3769869;rs79953491;rs355794;rs3820981 |
| 19 | 2:171631258:G:T | rs4668314 | 2 | 171631258 | 171570488 | 171680587 | 1.37E-08 | MetS | rs4668314 |

| | | | | | | | | | |
|---|---|---|---|---|---|---|---|---|---|
| 20 | 2:175241566:A:G | rs72917544 | 2 | 175241566 | 175236016 | 175241566 | 8.79E-10 | - | rs72917544 |
| 21 | 2:181607676:A:C | rs9630985 | 2 | 181607676 | 181518061 | 181618654 | 4.14E-10 | - | rs9630985;rs12615821 |
| 22 | 2:227006805:A:G | rs6724899 | 2 | 227006805 | 226813090 | 227123638 | 2.01E-14 | MetS; T2DM | rs6724899;rs2948565;rs2972151;rs1607365;rs1818829;rs4675045;rs57634424 |
| 23 | 2:228998742:A:G | rs72967047 | 2 | 228998742 | 228971784 | 229021719 | 5.55E-12 | - | rs72967047;rs4246656;rs13007222 |
| 24 | 2:230814719:C:T | rs6722477 | 2 | 230814719 | 230616197 | 230854290 | 4.56E-08 | - | rs6722477 |
| 25 | 3:12108754:C:T | rs11716141 | 3 | 12108754 | 12108754 | 12250888 | 1.11E-09 | T2DM | rs11716141 |
| 26 | 3:15718652:A:T | rs2470540 | 3 | 15718652 | 15680525 | 15970334 | 7.74E-10 | - | rs2470540;rs2470521 |
| 27 | 3:50041313:C:T | rs6765484 | 3 | 50041313 | 49669948 | 50248954 | 1.11E-15 | MetS; T2DM; ADHD; Anorexia; MDD | rs6765484;rs2624847;rs1046953;rs2518795;rs7648987;rs13316065;rs2280405;rs7613360;rs4688760 |
| 28 | 3:53125585:C:T | rs2564921 | 3 | 53125585 | 52970877 | 53139977 | 9.50E-09 | T2DM | rs2564921 |
| 29 | 3:62459819:A:C | rs76824303 | 3 | 62459819 | 62459819 | 62459819 | 3.43E-08 | - | rs76824303 |
| 30 | 3:63897215:C:T | rs2292662 | 3 | 63897215 | 63853423 | 64007214 | 3.63E-11 | T2DM | rs2292662;rs56030924 |
| 31 | 3:94038085:C:G | rs1454687 | 3 | 94038085 | 93883640 | 94198182 | 1.91E-10 | - | rs1454687;rs35319860 |
| 32 | 3:131632210:C:T | rs1228588 | 3 | 131632210 | 131447660 | 131792411 | 6.00E-11 | MetS | rs1228588;rs9289412 |
| 33 | 3:135955604:C:T | rs895893 | 3 | 135955604 | 135656190 | 136752590 | 2.54E-17 | MetS | rs895893;rs650505;rs6783337;rs9861150;rs1394094;rs2343681;rs6773440;rs148874568;rs9845457;rs13081352;rs9850364 |
| 34 | 3:150097635:C:G | rs9844972 | 3 | 150097635 | 150066540 | 150097635 | 2.65E-09 | T2DM | rs9844972 |
| 35 | 3:173117548:G:T | rs546738 | 3 | 173117548 | 173044559 | 173127808 | 4.18E-09 | - | rs546738 |
| 36 | 4:3241845:C:T | rs362307 | 4 | 3241845 | 3142660 | 3422211 | 1.17E-09 | T2DM | rs362307;rs12500459;rs10012797;rs3135063 |
| 37 | 4:45175691:C:T | rs13130484 | 4 | 45175691 | 45068929 | 45187622 | 3.26E-11 | T2DM | rs13130484;rs1849338 |
| 38 | 4:67802428:C:T | rs12509944 | 4 | 67802428 | 67802098 | 67903330 | 5.12E-09 | MetS | rs12509944 |
| 39 | 4:96170439:C:T | rs4699837 | 4 | 96170439 | 96123023 | 96181240 | 4.36E-08 | - | rs4699837 |
| 40 | 4:103679117:G:T | rs223490 | 4 | 103679117 | 103557311 | 104378012 | 1.37E-10 | T2DM | rs223490;rs10516497;rs13130741;rs7691873;rs1580278;rs58611096;rs227278;rs227372 |
| 41 | 4:137094048:A:C | rs1724557 | 4 | 137094048 | 136966779 | 137098470 | 9.24E-11 | - | rs1724557 |
| 42 | 4:140892872:C:T | rs1397440 | 4 | 140892872 | 140858717 | 140948095 | 4.02E-09 | - | rs1397440 |
| 43 | 5:53271420:A:G | rs702634 | 5 | 53271420 | 53271420 | 53287099 | 1.53E-09 | T2DM | rs702634 |
| 44 | 5:55861894:A:G | rs9687846 | 5 | 55861894 | 55794632 | 55876283 | 6.01E-16 | MetS; T2DM | rs30351;rs9687846;rs71624138;rs3900856 |
| 45 | 5:57607142:A:T | rs151912 | 5 | 57607142 | 57527886 | 57636145 | 3.23E-08 | - | rs151912 |

| | | | | | | | | | |
|---|---|---|---|---|---|---|---|---|---|
| 46 | 5:75003678:C:T | rs2307111 | 5 | 75003678 | 74568235 | 75038901 | 2.48E-15 | MetS; T2DM | rs2307111;rs13356670;rs34341;rs253413;rs11746063;rs888789 |
| 47 | 5:78522895:C:T | rs10078815 | 5 | 78522895 | 78416416 | 78628979 | 1.40E-10 | T2DM | rs10078815 |
| 48 | 5:87703099:C:G | rs7445169 | 5 | 87703099 | 87440497 | 88010829 | 7.28E-11 | ADHD; MDD | rs7445169;rs1477290 |
| 49 | 5:103995368:A:G | rs325485 | 5 | 103995368 | 103791044 | 104055261 | 2.88E-08 | ADHD; MDD | rs325485 |
| 50 | 5:118715275:G:T | rs924623 | 5 | 118715275 | 118697849 | 118800074 | 2.09E-08 | - | rs924623 |
| 51 | 6:7275468:C:T | rs9502575 | 6 | 7275468 | 7270087 | 7306153 | 2.30E-08 | T2DM | rs9502575 |
| 52 | 6:20518450:A:G | rs6903706 | 6 | 20518450 | 20483407 | 20632766 | 3.78E-10 | T2DM | rs6903706;rs2143407 |
| 53 | 6:26200677:A:G | rs806794 | 6 | 26200677 | 26180634 | 26259807 | 3.19E-09 | MDD | rs806794 |
| 54 | 6:33546498:C:T | rs5745582 | 6 | 33546498 | 33463961 | 33803752 | 1.77E-09 | - | rs5745582;rs72882008;rs9469485;rs210132;rs11755421;rs12055409 |
| 55 | 6:34749849:A:G | rs9689160 | 6 | 34749849 | 34549699 | 34832661 | 1.10E-08 | MetS | rs9689160 |
| 56 | 6:40369081:C:T | rs34045288 | 6 | 40369081 | 40348653 | 40383533 | 3.31E-08 | - | rs34045288 |
| 57 | 6:43757896:A:C | rs998584 | 6 | 43757896 | 43756863 | 43765533 | 2.01E-16 | MetS; T2DM | rs998584;rs6458349 |
| 58 | 6:50788778:A:C | rs3798519 | 6 | 50788778 | 50664180 | 50940677 | 3.41E-13 | T2DM | rs3798519;rs72889996;rs2206271;rs62405419 |
| 59 | 6:79349892:A:C | rs236852 | 6 | 79349892 | 79316046 | 79477484 | 4.24E-08 | - | rs236852 |
| 60 | 6:127454893:A:G | rs72959041 | 6 | 127454893 | 127439897 | 127529780 | 9.91E-09 | MetS | rs72959041 |
| 61 | 6:139835329:A:T | rs2982521 | 6 | 139835329 | 139828916 | 139844429 | 1.34E-09 | MetS | rs2982521 |
| 62 | 6:153371503:C:T | rs7761831 | 6 | 153371503 | 153350709 | 153449670 | 2.36E-10 | T2DM | rs7761831;rs17712040;rs6932473 |
| 63 | 7:1929019:G:T | rs73048106 | 7 | 1929019 | 1824028 | 2110850 | 8.71E-11 | MDD | rs73048106;rs55893771;rs55860148;rs7807190;rs7680 |
| 64 | 7:2760750:A:C | rs798549 | 7 | 2760750 | 2760750 | 2862891 | 2.72E-08 | - | rs798549 |
| 65 | 7:17287998:A:G | rs2106727 | 7 | 17287998 | 17281007 | 17309279 | 9.10E-09 | - | rs2106727 |
| 66 | 7:28210660:C:T | rs520161 | 7 | 28210660 | 28173522 | 28214614 | 1.50E-08 | T2DM | rs520161 |
| 67 | 7:44255643:A:G | rs878521 | 7 | 44255643 | 44223721 | 44265179 | 5.47E-10 | T2DM | rs878521 |
| 68 | 7:50683678:A:G | rs2715126 | 7 | 50683678 | 50659193 | 50685625 | 5.91E-09 | - | rs2715126 |
| 69 | 7:130466091:A:G | rs1447060 | 7 | 130466091 | 130422934 | 130468190 | 3.89E-13 | MetS; T2DM | rs1447060;rs11762784;rs10260148 |
| 70 | 7:150657095:A:G | rs56282717 | 7 | 150657095 | 150610935 | 150660239 | 8.23E-10 | MetS | rs56282717 |
| 71 | 8:9183358:A:G | rs9987289 | 8 | 9183358 | 9177732 | 9231661 | 1.38E-11 | MetS; T2DM | rs9987289;rs7357361 |
| 72 | 8:9861116:C:T | rs6601406 | 8 | 9861116 | 9860588 | 10005526 | 8.21E-10 | T2DM | rs6601406;rs17689007 |

| | | | | | | | | | |
|---|---|---|---|---|---|---|---|---|---|
| 73 | 8:10675500:C:G | rs2409656 | 8 | 10675500 | 10570877 | 10708509 | 5.19E-09 | T2DM | rs2409656 |
| 74 | 8:19955382:C:T | rs113149294 | 8 | 19955382 | 19526176 | 19974789 | 4.27E-18 | MetS; T2DM | rs17091237;rs7836144;rs58558359;rs6586863;rs13282247;rs2035891;rs1472165;rs10105881;rs56225720;rs28585593;rs60329942;rs3927029;rs6998248;rs11204079;rs10503660;rs17410086;rs11781244;rs28413168;rs78226669;rs1031046;rs142084074;rs111648015;rs1534649;rs76450127;rs308;rs2165557;rs35014088;rs75902744;rs186026600;rs117174179;rs28630415;rs7011846;rs1470186;rs276;rs73208821;rs113149294;rs74737417 |
| 75 | 8:30863938:C:T | rs10954772 | 8 | 30863938 | 30819854 | 30879207 | 3.12E-12 | MetS | rs10954772;rs11775287;rs2543617 |
| 76 | 8:116560619:C:T | rs7825679 | 8 | 116560619 | 116409058 | 116645056 | 9.66E-13 | MetS; T2DM | rs7825679;rs2737226;rs800899 |
| 77 | 8:126631139:A:T | rs1551393 | 8 | 126631139 | 126628841 | 126648243 | 9.71E-09 | MetS | rs1551393 |
| 78 | 9:28410683:C:T | rs1412234 | 9 | 28410683 | 28410683 | 28488365 | 2.51E-09 | T2DM | rs1412234 |
| 79 | 9:134849037:A:G | rs11243586 | 9 | 134849037 | 134802170 | 134946805 | 1.00E-08 | - | rs11243586 |
| 80 | 10:12297230:C:T | rs4747969 | 10 | 12297230 | 12245520 | 12328010 | 5.54E-10 | T2DM | rs4747969;rs7069060 |
| 81 | 10:64988931:A:T | rs7916868 | 10 | 64988931 | 64874754 | 65400352 | 2.63E-12 | MetS | rs7916868;rs111346856;rs10995566;rs3858122;rs4564213 |
| 82 | 10:80985374:A:G | rs1250591 | 10 | 80985374 | 80906729 | 81067040 | 2.39E-13 | T2DM | rs703994;rs1250591;rs2802363;rs1104906;rs1250595;rs1749850;rs1250575;rs942796;rs1250573 |
| 83 | 10:94138239:A:G | rs7903767 | 10 | 94138239 | 93544818 | 94500111 | 1.42E-13 | T2DM | rs7903767;rs2995787;rs7084673;rs2901587;rs7894946;rs1776209;rs1099379;rs2251101;rs117704022;rs11187152 |
| 84 | 10:99772404:A:G | rs563296 | 10 | 99772404 | 99762693 | 99784552 | 5.25E-09 | MetS | rs563296 |
| 85 | 10:113940329:C:T | rs2792751 | 10 | 113940329 | 113901194 | 113950418 | 1.65E-08 | - | rs2792751 |
| 86 | 11:2854514:C:G | rs234862 | 11 | 2854514 | 2844216 | 2854514 | 2.13E-08 | T2DM | rs234862 |
| 87 | 11:8694830:A:G | rs10128597 | 11 | 8694830 | 8440969 | 8694830 | 2.56E-09 | - | rs10128597 |
| 88 | 11:13269921:A:T | rs11825504 | 11 | 13269921 | 13268386 | 13350131 | 1.56E-08 | MetS | rs11825504;rs11824092 |
| 89 | 11:27722298:G:T | rs7944119 | 11 | 27722298 | 27477864 | 27748493 | 3.65E-12 | MetS | rs7944119;rs12273363;rs11030075;rs12276130 |
| 90 | 11:43752522:A:G | rs10838158 | 11 | 43752522 | 43634973 | 43878534 | 9.39E-09 | T2DM | rs10838158 |
| 91 | 11:47529947:A:C | rs7124681 | 11 | 47529947 | 47232038 | 47946836 | 9.43E-16 | MetS; T2DM | rs7124681;rs7950674;rs1685404;rs11570094 |
| 92 | 11:57268252:C:T | rs112450479 | 11 | 57268252 | 57158320 | 57386178 | 4.73E-08 | - | rs112450479 |
| 93 | 11:65662752:A:G | rs588114 | 11 | 65662752 | 65211979 | 65663547 | 2.63E-10 | MetS; T2DM | rs3825071;rs1346;rs10750766;rs588114 |
| 94 | 11:76485358:A:G | rs737185 | 11 | 76485358 | 76464812 | 76511271 | 7.51E-09 | - | rs737185 |
| 95 | 11:102948592:A:G | rs2510087 | 11 | 102948592 | 102929625 | 103153310 | 2.64E-08 | - | rs2510087 |

| | | | | | | | | | |
|---|---|---|---|---|---|---|---|---|---|
| 96 | 11:116675294:A:T | rs6589570 | 11 | 116675294 | 116527728 | 117186506 | 5.06E-13 | MetS | rs2367970;rs1263151;rs6589570;rs7950633;rs1729408;rs2849174;rs116987336 |
| 97 | 11:118949083:A:G | rs1177563 | 11 | 118949083 | 118913993 | 118958869 | 5.58E-10 | - | rs1177563 |
| 98 | 12:26320656:C:T | rs10842685 | 12 | 26320656 | 26313849 | 26339986 | 2.70E-08 | T2DM | rs10842685 |
| 99 | 12:27919145:C:T | rs9668691 | 12 | 27919145 | 27905969 | 27965150 | 9.80E-09 | T2DM | rs9668691 |
| 100 | 12:50263148:A:G | rs7132908 | 12 | 50263148 | 50240304 | 50285780 | 1.62E-10 | MetS | rs7132908;rs7305229 |
| 101 | 12:56467865:C:G | rs10876869 | 12 | 56467865 | 56368078 | 56609885 | 9.06E-09 | - | rs10876869 |
| 102 | 12:108618630:C:T | rs3764002 | 12 | 108618630 | 108609634 | 108629780 | 1.65E-10 | T2DM | rs3764002 |
| 103 | 12:123447928:C:T | rs4275659 | 12 | 123447928 | 123296204 | 123913433 | 1.30E-10 | T2DM | rs4275659;rs12820906;rs4930719;rs138062324 |
| 104 | 12:124409502:A:G | rs7133378 | 12 | 124409502 | 124401710 | 124496316 | 5.27E-09 | - | rs7133378 |
| 105 | 13:58628636:C:T | rs8000972 | 13 | 58628636 | 58254773 | 58828747 | 2.64E-10 | T2DM | rs8000972;rs9569820 |
| 106 | 13:80644618:A:G | rs12431307 | 13 | 80644618 | 80620642 | 80758943 | 1.25E-08 | T2DM | rs12431307;rs17804744 |
| 107 | 14:23823913:A:T | rs7148564 | 14 | 23823913 | 23792416 | 23837484 | 3.26E-08 | - | rs7148564 |
| 108 | 14:25930988:A:C | rs8015400 | 14 | 25930988 | 25927298 | 25994917 | 2.99E-08 | - | rs8015400 |
| 109 | 14:79891882:A:G | rs10150482 | 14 | 79891882 | 79833494 | 79945162 | 4.58E-10 | T2DM | rs10150482 |
| 110 | 15:41855736:G:T | rs1023193 | 15 | 41855736 | 41798762 | 42190692 | 1.90E-11 | MetS; T2DM | rs1023193;rs2277536;rs12438252;rs890503;rs2254078;rs1200346;rs9302110 |
| 111 | 15:51893161:C:T | rs4774595 | 15 | 51893161 | 51709536 | 52004250 | 3.78E-09 | - | rs4774595 |
| 112 | 15:56870157:C:T | rs139280016 | 15 | 56870157 | 56825676 | 57633098 | 4.93E-10 | T2DM | rs139280016;rs139219993;rs12439266 |
| 113 | 15:58678720:C:T | rs261290 | 15 | 58678720 | 58671559 | 58741891 | 3.37E-16 | MetS | rs261290;rs75437656;rs28698356;rs723967;rs12438999;rs11854624;rs1601935;rs12908474;rs261334 |
| 114 | 15:63922474:C:T | rs11071759 | 15 | 63922474 | 63798569 | 64136472 | 1.05E-09 | T2DM | rs11071759;rs7178762 |
| 115 | 15:68080886:A:T | rs4776970 | 15 | 68080886 | 67848128 | 68113771 | 1.48E-09 | T2DM | rs4776970 |
| 116 | 15:73451443:A:C | rs1904335 | 15 | 73451443 | 73392760 | 73621435 | 1.34E-09 | - | rs1904335 |
| 117 | 15:77799657:A:G | rs4886869 | 15 | 77799657 | 77310345 | 77912717 | 5.40E-15 | T2DM | rs35268067;rs11631927;rs8042307;rs2271396;rs62008430;rs11852471;rs4886869;rs939486;rs35134156 |
| 118 | 16:387037:A:C | rs7498932 | 16 | 387037 | 376836 | 412974 | 2.29E-08 | - | rs7498932 |
| 119 | 16:24735736:C:T | rs200538 | 16 | 24735736 | 24709089 | 24832693 | 2.05E-08 | MetS | rs200538;rs7188873 |
| 120 | 16:28917644:C:T | rs7188071 | 16 | 28917644 | 28403471 | 28955702 | 3.29E-09 | - | rs7188071 |
| 121 | 16:30018720:C:T | rs12921753 | 16 | 30018720 | 29923510 | 30147265 | 4.32E-15 | MetS; T2DM | rs12921753;rs7542;rs8059619;rs12928610 |

| | | | | | | | | | |
|---|---|---|---|---|---|---|---|---|---|
| 122 | 16:31131174:A:G | rs4889620 | 16 | 31131174 | 30916233 | 31149142 | 1.53E-11 | Obesity | rs4889620 |
| 123 | 16:53843533:C:T | rs6499646 | 16 | 53843533 | 53805344 | 53843533 | 1.96E-08 | MetS; Obesity; T2DM | rs16952522;rs6499646 |
| 124 | 16:57001985:A:C | rs60545348 | 16 | 57001985 | 56898198 | 57006305 | 7.30E-10 | MetS | rs1968493;rs60545348 |
| 125 | 16:69651866:C:T | rs862320 | 16 | 69651866 | 69547741 | 70112729 | 1.75E-14 | MetS; T2DM | rs862320;rs4275849;rs4985376;rs9746247;rs904804;rs11866219;rs62052820 |
| 126 | 16:81534790:C:T | rs2925979 | 16 | 81534790 | 81449060 | 81549708 | 1.18E-15 | MetS; T2DM | rs2550733;rs35081994;rs2925979;rs2927307;rs1966957;rs2966085;rs66710707 |
| 127 | 17:1830836:C:T | rs77483079 | 17 | 1830836 | 1820080 | 1859542 | 1.25E-09 | - | rs77483079;rs4790849 |
| 128 | 17:3882309:A:G | rs17763551 | 17 | 3882309 | 3880546 | 4165741 | 3.58E-11 | T2DM | rs17763551;rs8068804;rs36019144 |
| 129 | 17:29720140:C:T | rs8065496 | 17 | 29720140 | 29470288 | 29735829 | 6.15E-09 | - | rs8065496 |
| 130 | 17:34850623:G:T | rs7222903 | 17 | 34850623 | 34825861 | 34961772 | 5.00E-09 | - | rs7222903;rs34874411 |
| 131 | 17:40721042:C:T | rs650558 | 17 | 40721042 | 40565200 | 41018481 | 6.22E-12 | MetS; T2DM | rs650558;rs72628307 |
| 132 | 17:42096209:C:T | rs77034317 | 17 | 42096209 | 41952807 | 42282225 | 1.35E-08 | MetS | rs231492;rs77034317 |
| 133 | 17:46124326:C:G | rs9900074 | 17 | 46124326 | 45986496 | 46323934 | 7.71E-09 | - | rs9900074 |
| 134 | 17:47090785:C:T | rs11079849 | 17 | 47090785 | 46835629 | 47145848 | 2.40E-11 | T2DM | rs11079849;rs11654075;rs55917610;rs12601955;rs4794008;rs55797392 |
| 135 | 17:65832576:A:G | rs12452511 | 17 | 65832576 | 65764164 | 66096529 | 9.23E-13 | MetS; T2DM | rs12452511;rs62086892;rs11650220;rs72631329 |
| 136 | 17:76405736:C:T | rs4969143 | 17 | 76405736 | 76386037 | 76406170 | 1.57E-08 | MetS | rs4969143 |
| 137 | 17:76798155:C:T | rs2306527 | 17 | 76798155 | 76661269 | 76833916 | 2.69E-08 | - | rs2306527 |
| 138 | 18:1839911:G:T | rs60764613 | 18 | 1839911 | 1811604 | 1914051 | 2.76E-08 | - | rs60764613 |
| 139 | 18:21134239:C:T | rs1631685 | 18 | 21134239 | 21074255 | 21165409 | 7.39E-11 | MetS | rs1631685;rs1788784 |
| 140 | 18:57848651:A:G | rs66922415 | 18 | 57848651 | 57728947 | 58090217 | 1.01E-20 | MetS; Obesity; T2DM | rs66922415;rs9947403;rs489693;rs79077117;rs4121765;rs1942866;rs11660783;rs487720;rs73455661;rs10503040;rs58067248;rs1943226;rs78138914;rs17066842;rs8086205 |
| 141 | 19:7970635:A:G | rs4804833 | 19 | 7970635 | 7931242 | 7978430 | 3.59E-11 | MetS; T2DM | rs572840;rs2115108;rs4804833 |
| 142 | 19:8432737:C:T | rs117760119 | 19 | 8432737 | 8432737 | 8441959 | 1.57E-08 | MetS | rs117760119 |
| 143 | 19:33890838:C:G | rs10406327 | 19 | 33890838 | 33890838 | 33897478 | 1.97E-08 | T2DM | rs10406327 |
| 144 | 19:46157237:A:G | rs10407429 | 19 | 46157237 | 46108455 | 46201213 | 4.33E-13 | T2DM | rs10407429;rs10409882;rs34078197;rs2334253;rs35650950 |
| 145 | 19:47569003:A:G | rs3810291 | 19 | 47569003 | 47559074 | 47623080 | 1.71E-14 | MetS; T2DM | rs3810291 |
| 146 | 20:32657378:C:T | rs932388 | 20 | 32657378 | 32514061 | 32729444 | 1.90E-08 | T2DM | rs932388 |

| | | | | | | | | | |
|---|---|---|---|---|---|---|---|---|---|
| 147 | 20:43042364:C:T | rs1800961 | 20 | 43042364 | 42958768 | 43042364 | 1.32E-16 | MetS; T2DM | rs1800961;rs150224153 |
| 148 | 20:50986299:A:G | rs13037010 | 20 | 50986299 | 50809213 | 51264331 | 1.84E-12 | MetS | rs13037010;rs6068177;rs74687627;rs2143901;rs6068280;rs13042290;rs6021835;rs6021886 |
| 149 | 20:62450664:C:T | rs6011155 | 20 | 62450664 | 62402860 | 62471527 | 2.21E-08 | - | rs6011155 |
| 150 | 21:46494995:C:T | rs9977825 | 21 | 46494995 | 46487831 | 46558780 | 4.32E-08 | - | rs9977825 |

Note. Genomic risk loci were identified using FUMA (v1.5.6; Watanabe et al 2017). BP refers to the base pair position of the index SNP. Start and End columns refer to the BP positions of the borders of the associated genomic loci. All genome-wide significant independent (at $r2 < 0.6$) SNPs within each locus are listed in the last column. Genomic risk loci overlap was assessed by comparing the start and end sites of each loci with those indicated by FUMA with the univariate GWAS summary statistics contributing to the Psych-IR factor.

**Supplementary Tables S7-S16. In the excel file.**

**Supplementary Figures**

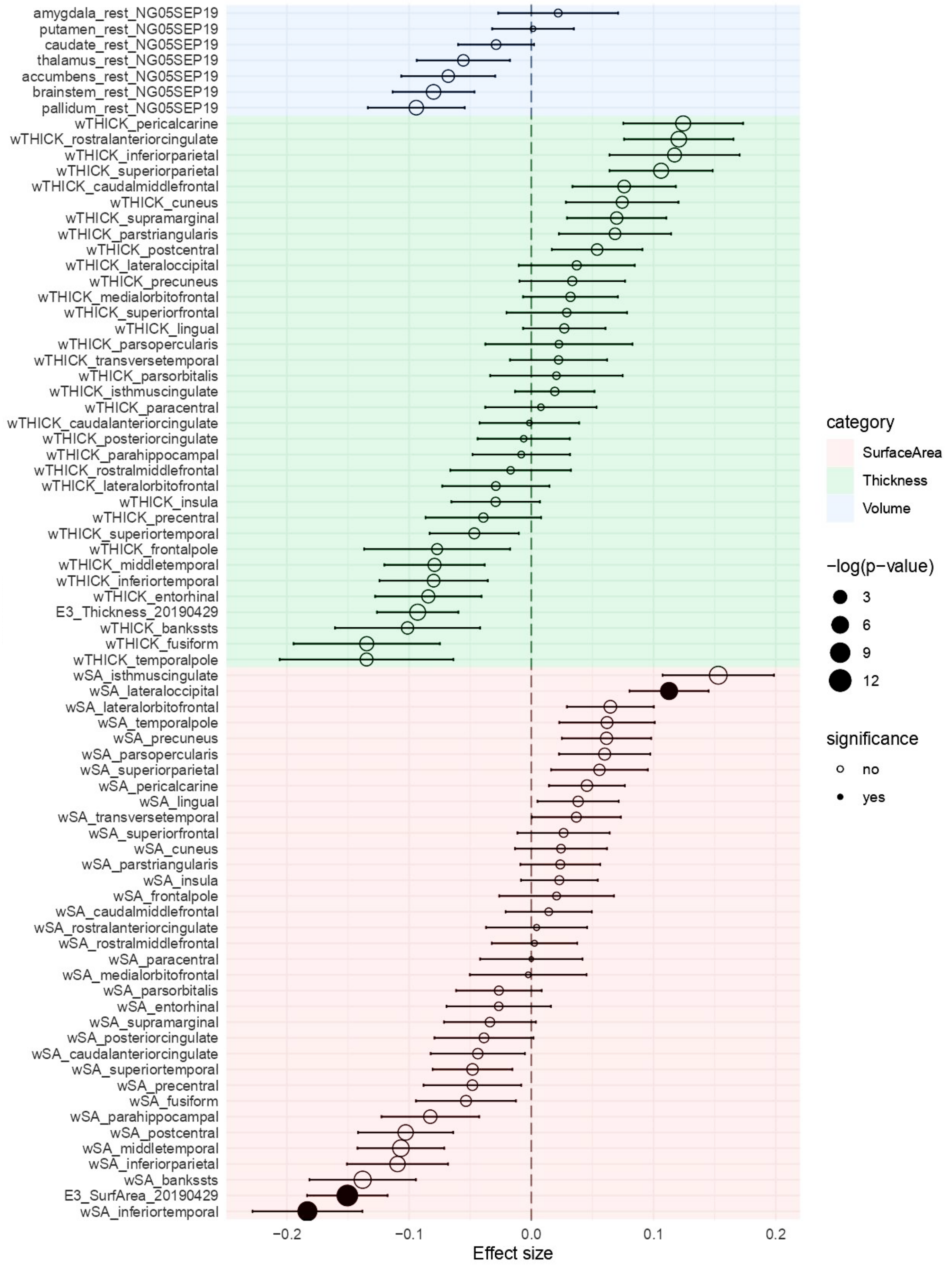

**Supplementary Figure S1. Genetic correlation estimates between brain morphometric traits and the Psych-IR multimorbidity factor.**

Background color distinguishes subcortical volumes (blue), cortical thickness (green) and surface area (red). For each brain trait, the circle represents the point estimate of genetic correlation and the circle size the strength of association. Filled circles are significant genetic correlations when considering the Bonferroni significance threshold of $P_{Bonf}$=6.41E-04 (i.e. 0.05/78 brain traits tested).

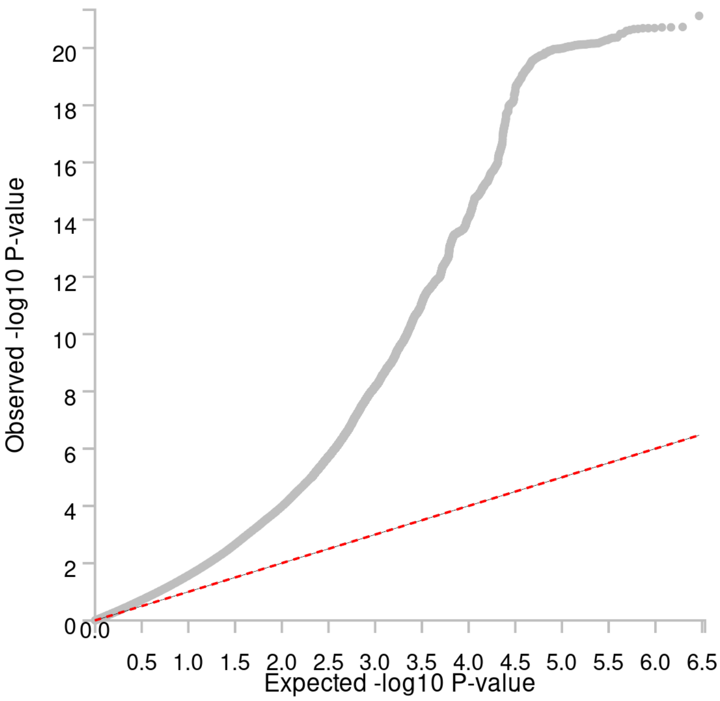

**Supplementary Figure S2. Quantile-quantile plot of the -log10 P-values from the Psych-IR multimorbidity factor multivariate GWAS, after the removal of significant Q-SNPs (and those in LD with Q-SNPs).**

The red line indicates the distribution of P-values under the null hypothesis of no association.

a)

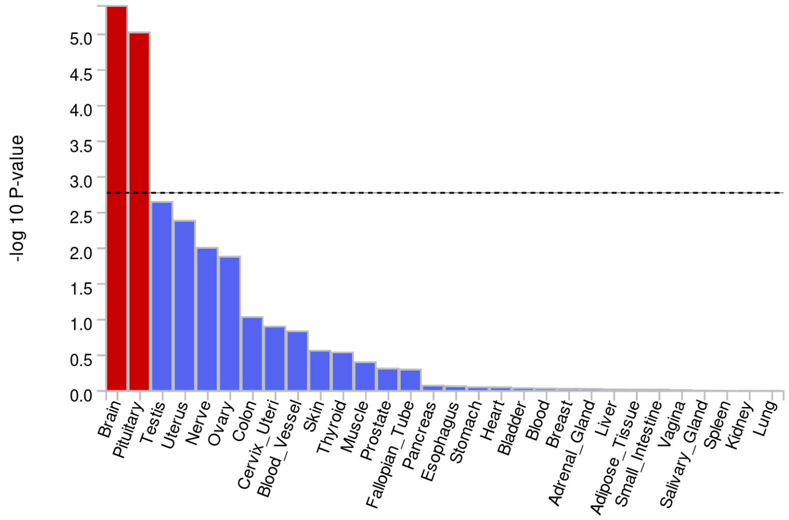

b)

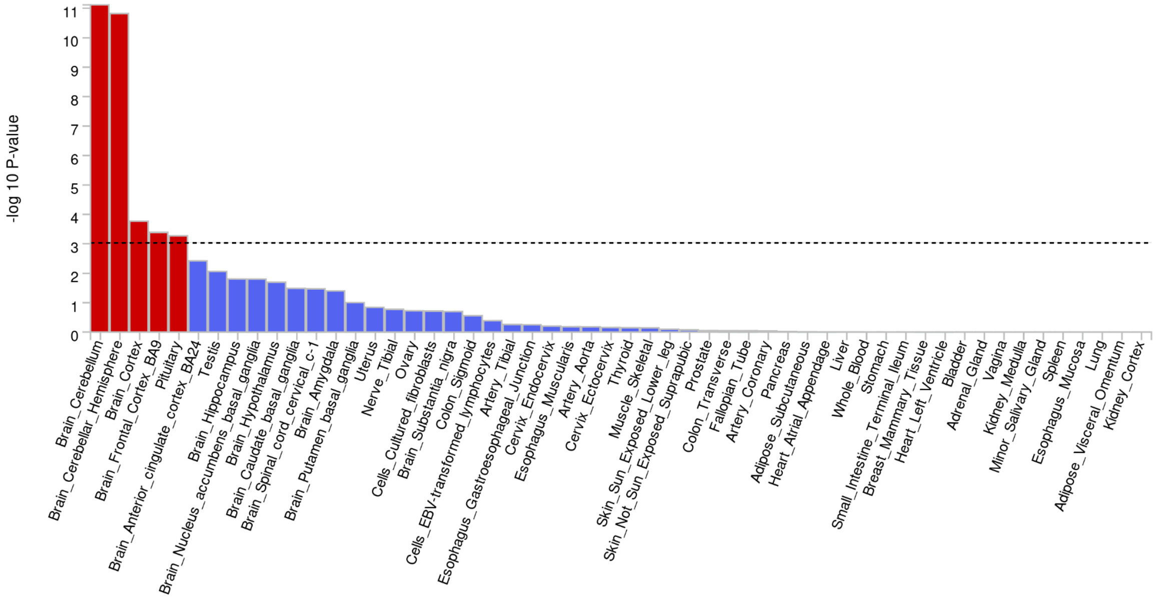

c)

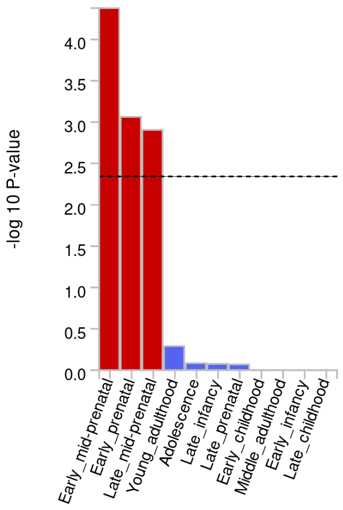

**Supplementary Figure S3. Gene-property analyses results.**

Figure shows the relationship between the Psych-IR multimorbidity factor gene-based associations and tissue specific gene expression profiles across a) 30 general tissues types (GTEx v8), b) 54 tissues (GTEx v8), and c) 11 developmental stages of the brain (BrainSpan). Red bars denote significant tissues or developmental stages.

Top 6 genes by tissue

Z-scores coloured by intensity

| Tissue | Gene_top1 | Gene_top2 | Gene_top3 | Gene_top4 | Gene_top5 | Gene_top6 | Z_Est_top1 | Z_Est_top2 | Z_Est_top3 | Z_Est_top4 | Z_Est_top5 | Z_Est_top6 |
|---|---|---|---|---|---|---|---|---|---|---|---|---|
| Brain_Amygdala | RP11-69E11.4 | MTCH2 | SNF8 | RNF123 | RPAP1 | KAT8 | 10.745 | 7.730 | 7.583 | 7.451 | -7.079 | 7.078 |
| Brain_Anterior_cingulate_cortex_BA24 | RBM6 | INO80E | MST1R | MTCH2 | RNF123 | MAPK3 | -9.145 | -9.089 | 9.081 | 7.748 | 7.727 | -7.275 |
| Brain_Caudate_basal_ganglia | RBM6 | UBA7 | LINGO1 | AC009133.12 | RP11-69E11.4 | INO80E | -9.126 | -8.874 | 8.338 | 8.263 | 8.207 | -8.138 |
| Brain_Cerebellar_Hemisphere | RBM6 | PCCB | C1QTNF4 | NR1H3 | NDUFS3 | INO80E | -9.182 | 9.162 | -9.085 | 9.084 | 8.908 | -8.848 |
| Brain_Cerebellum | MST1R | C1QTNF4 | RBM6 | INO80E | NDUFS3 | PROX1-AS1 | 9.295 | -9.291 | -9.099 | -8.697 | 7.888 | 7.619 |
| Brain_Cortex | RBM6 | MST1R | SLX1B | INO80E | BMP8A | RAPSN | -9.200 | 8.804 | 8.738 | -8.600 | 8.538 | -8.284 |
| Brain_Frontal_Cortex_BA9 | RBM6 | INO80E | BMP8A | OXCT2P1 | LINC00674 | RP11-455F5.3 | -9.215 | -8.801 | 8.770 | 8.682 | 7.949 | -7.634 |
| Brain_Hippocampus | RBM6 | LINGO1 | PROX1-AS1 | BMP8A | INO80E | PPIEL | -9.158 | 8.735 | 8.692 | 8.549 | -8.370 | 8.180 |
| Brain_Hypothalamus | BMP8A | RBM6 | LINGO1-AS1 | MST1R | OXCT2P1 | RNF123 | 11.413 | -9.232 | 8.698 | 8.567 | 8.510 | 8.340 |
| Brain_Nucleus_accumbens_basal_ganglia | RP11-69E11.4 | RP11-416A14.1 | MST1R | RBM6 | TMEM219 | RNF123 | 10.676 | -9.976 | 9.210 | -9.209 | -8.695 | 7.451 |
| Brain_Putamen_basal_ganglia | RBM6 | LINGO1 | INO80E | RP11-69E11.4 | MTCH2 | LINGO1-AS1 | -9.191 | 8.998 | -8.827 | 8.527 | 7.345 | 7.302 |
| Brain_Spinal_cord_cervical_c-1 | RP11-69E11.4 | BMP8A | RBM6 | INO80E | MST1R | LINGO1-AS1 | 11.251 | 9.852 | -9.023 | -8.842 | 8.430 | 8.073 |
| Brain_Substantia_nigra | RBM6 | RP11-478J18.2 | RP11-307C19.1 | KAT8 | C12orf65 | JMJD7 | -9.085 | 7.821 | 7.201 | 7.101 | 7.050 | 7.004 |
| PEC_Prefrontal_cortex | MACF1 | HPCAL4 | NOB1 | MTCH2 | KBTBD4 | CELF1 | 11.217 | 8.759 | -8.653 | 7.478 | 7.103 | -7.047 |
| Pituitary | RP11-420K8.1 | RBM6 | RNF123 | INO80E | PABPC4 | RP11-731C17.2 | 9.909 | -9.217 | 9.184 | -8.700 | -8.208 | 7.329 |

**Supplementary Figure S4. Heatmap of the top six most significant genes by tissue, as identified by Transcriptome-wide Structural Equation Modeling (T-SEM), excluding the MHC region.**

The figure is organized with the gene names listed on the left side, while their corresponding Z-scores are shown on the right side. The heatmap uses color intensity to reflect the magnitude of Z-scores, with warmer tones (orange) indicating higher positive associations and cooler tones (blue) representing negative associations. Each row corresponds to a brain region.

### Top 16 most significant genes across all tissues

Cells are coloured according to Z-scores only for tissues where specific gene expression association was found

| Tissue | BMP8A | RP11-69E11.4 | MACF1 | RP11-416A14.1 | RP11-420K8.1 | MST1R | C1QTNF4 | RBM6 | RNF123 | PCCB | INO80E | NR1H3 | LINGO1 | NDUFS3 | UBA7 | HPCAL4 |
|---|---|---|---|---|---|---|---|---|---|---|---|---|---|---|---|---|
| Brain_Amygdala | - | 10.745278 | - | - | - | - | - | -4.698159 | 7.450906 | - | - | - | - | - | - | - |
| Brain_Anterior_cingulate_cortex_BA24 | - | - | - | - | - | 9.080789 | - | -9.145329 | 7.727368 | - | -9.089012 | - | - | - | - | - |
| Brain_Caudate_basal_ganglia | - | 8.207103 | - | - | - | 7.597018 | -6.920960 | -9.125929 | - | - | -8.138074 | - | 8.338361 | - | -8.874415 | 5.702499 |
| Brain_Cerebellar_Hemisphere | - | 5.939722 | - | - | - | 8.529004 | -9.084876 | -9.182047 | 7.503368 | 9.161730 | -8.847738 | 9.084418 | - | 8.907613 | - | - |
| Brain_Cerebellum | - | 6.232011 | - | - | - | 9.294794 | -9.290762 | -9.098548 | 7.292161 | 5.803149 | -8.697477 | - | - | 7.888057 | - | - |
| Brain_Cortex | 8.538309 | 7.694717 | - | - | - | 8.804225 | - | -9.200340 | 8.175184 | - | -8.600185 | - | 5.781797 | - | - | - |
| Brain_Frontal_Cortex_BA9 | 8.769599 | 6.594851 | - | - | - | 7.463174 | - | -9.215190 | 7.449576 | 6.433872 | -8.801158 | - | - | - | - | - |
| Brain_Hippocampus | 8.548722 | 7.388470 | - | - | - | - | -6.495978 | -9.157651 | - | - | -8.370329 | - | 8.734676 | - | - | - |
| Brain_Hypothalamus | 11.412795 | 7.787702 | - | - | - | 8.567151 | - | -9.231757 | 8.339620 | - | -5.634188 | - | 6.574690 | - | - | - |
| Brain_Nucleus_accumbens_basal_ganglia | - | 10.676323 | - | -9.975992 | - | 9.210190 | - | -9.209289 | 7.451053 | - | - | - | 7.115560 | 7.332139 | - | 5.595877 |
| Brain_Putamen_basal_ganglia | - | 8.527109 | - | - | - | - | - | -9.191287 | 6.521591 | - | -8.826649 | - | 8.998337 | - | - | 5.469942 |
| Brain_Spinal_cord_cervical_c-1 | 9.852007 | 11.251321 | - | - | - | 8.429930 | - | -9.023353 | 7.873174 | - | -8.841851 | - | 5.079897 | - | - | - |
| Brain_Substantia_nigra | - | 5.872710 | - | - | - | - | - | -9.085172 | - | - | - | - | - | - | - | - |
| Pituitary | - | 7.269468 | - | - | 9.909219 | - | - | -9.217086 | 9.184165 | - | -8.699604 | - | - | - | - | 5.449284 |
| PEC_Prefrontal_cortex | - | - | 11.21723 | - | - | - | - | - | - | - | -6.173981 | - | - | - | - | 8.758629 |

**Supplementary Figure S5. Heatmap of the top 16 most significant genes across multiple brain tissues identified via Transcriptome-wide Structural Equation Modeling (T-SEM), excluding the MHC region.**

The heatmap illustrates tissue-specific gene expression associations, with the cells colored according to Z-scores, indicating the strength of these associations. Warmer colors (orange) correspond to higher positive Z-scores, indicating stronger positive associations, while cooler colors (blue) indicate negative Z-scores. Each row represents a brain region or tissue.

Top 6 genes by tissue

Z-scores coloured by intensity

| Tissue | Gene_top1 | Gene_top2 | Gene_top3 | Gene_top4 | Gene_top5 | Gene_top6 | Z_Est_top1 | Z_Est_top2 | Z_Est_top3 | Z_Est_top4 | Z_Est_top5 | Z_Est_top6 |
|---|---|---|---|---|---|---|---|---|---|---|---|---|
| Brain_Amygdala | RP11-69E11.4 | MTCH2 | SNF8 | RNF123 | HCG27 | RPAP1 | 10.745 | 7.730 | 7.583 | 7.451 | 7.149 | -7.079 |
| Brain_Anterior_cingulate_cortex_BA24 | RBM6 | INO80E | MST1R | HCG27 | MTCH2 | RNF123 | -9.145 | -9.089 | 9.081 | 7.966 | 7.748 | 7.727 |
| Brain_Caudate_basal_ganglia | RBM6 | UBA7 | LINGO1 | AC009133.12 | RP11-69E11.4 | INO80E | -9.126 | -8.874 | 8.338 | 8.263 | 8.207 | -8.138 |
| Brain_Cerebellar_Hemisphere | RBM6 | PCCB | C1QTNF4 | NR1H3 | NDUFS3 | INO80E | -9.182 | 9.162 | -9.085 | 9.084 | 8.908 | -8.848 |
| Brain_Cerebellum | MST1R | C1QTNF4 | RBM6 | INO80E | NOTCH4 | NDUFS3 | 9.295 | -9.291 | -9.099 | -8.697 | -8.528 | 7.888 |
| Brain_Cortex | RBM6 | MST1R | SLX1B | INO80E | BMP8A | RAPSN | -9.200 | 8.804 | 8.738 | -8.600 | 8.538 | -8.284 |
| Brain_Frontal_Cortex_BA9 | RBM6 | INO80E | BMP8A | OXCT2P1 | LINC00674 | RP11-455F5.3 | -9.215 | -8.801 | 8.770 | 8.682 | 7.949 | -7.634 |
| Brain_Hippocampus | RBM6 | LINGO1 | PROX1-AS1 | NOTCH4 | BMP8A | INO80E | -9.158 | 8.735 | 8.692 | -8.643 | 8.549 | -8.370 |
| Brain_Hypothalamus | BMP8A | RBM6 | LINGO1-AS1 | MST1R | OXCT2P1 | RNF123 | 11.413 | -9.232 | 8.698 | 8.567 | 8.510 | 8.340 |
| Brain_Nucleus_accumbens_basal_ganglia | RP11-69E11.4 | RP11-416A14.1 | MST1R | RBM6 | TMEM219 | HLA-DRB1 | 10.676 | -9.976 | 9.210 | -9.209 | -8.695 | -8.013 |
| Brain_Putamen_basal_ganglia | RBM6 | LINGO1 | INO80E | RP11-69E11.4 | CYP21A2 | MTCH2 | -9.191 | 8.998 | -8.827 | 8.527 | 7.433 | 7.345 |
| Brain_Spinal_cord_cervical_c-1 | RP11-69E11.4 | BMP8A | RBM6 | INO80E | MST1R | LINGO1-AS1 | 11.251 | 9.852 | -9.023 | -8.842 | 8.430 | 8.073 |
| Brain_Substantia_nigra | RBM6 | RP11-478J18.2 | RP11-307C19.1 | KAT8 | C12orf65 | LEMD2 | -9.085 | 7.821 | 7.201 | 7.101 | 7.050 | -7.018 |
| PEC_Prefrontal_cortex | MACF1 | HPCAL4 | NOB1 | MTCH2 | KBTBD4 | CELF1 | 11.217 | 8.759 | -8.653 | 7.478 | 7.103 | -7.047 |
| Pituitary | RP11-420K8.1 | RBM6 | RNF123 | INO80E | PABPC4 | RP11-731C17.2 | 9.909 | -9.217 | 9.184 | -8.700 | -8.208 | 7.329 |

**Supplementary Figure S6. Heatmap of the top six most significant genes by tissue, as identified by Transcriptome-wide Structural Equation Modeling (T-SEM), including the MHC region.**

The figure is organized with the gene names listed on the left side, while their corresponding Z-scores are shown on the right side. The heatmap uses color intensity to reflect the magnitude of Z-scores, with warmer tones (orange) indicating higher positive associations and cooler tones (blue) representing negative associations. Each row corresponds to a brain region.

Top 16 most significant genes across all tissues

Cells are coloured according to Z-scores only for tissues where specific gene expression association was found

| Tissue | BMP8A | RP11-69E11.4 | MACF1 | RP11-416A14.1 | RP11-420K8.1 | MST1R | C1QTNF4 | RBM6 | RNF123 | PCCB | INO80E | NR1H3 | LINGO1 | NDUFS3 | UBA7 | HPCAL4 |
|---|---|---|---|---|---|---|---|---|---|---|---|---|---|---|---|---|
| Brain_Amygdala | - | 10.745278 | - | - | - | - | - | -4.698159 | 7.450906 | - | - | - | - | - | - | - |
| Brain_Anterior_cingulate_cortex_BA24 | - | - | - | - | - | 9.080789 | - | -9.145329 | 7.727368 | - | -9.089012 | - | - | - | - | - |
| Brain_Caudate_basal_ganglia | - | 8.207103 | - | - | - | 7.597018 | -6.920960 | -9.125929 | - | - | -8.138074 | - | 8.338361 | - | -8.874415 | 5.702499 |
| Brain_Cerebellar_Hemisphere | - | 5.939722 | - | - | - | 8.529004 | -9.084876 | -9.182047 | 7.503368 | 9.161730 | -8.847738 | 9.084418 | - | 8.907613 | - | - |
| Brain_Cerebellum | - | 6.232011 | - | - | - | 9.294794 | -9.290762 | -9.098548 | 7.292161 | 5.803149 | -8.697477 | - | - | 7.888057 | - | - |
| Brain_Cortex | 8.538309 | 7.694717 | - | - | - | 8.804225 | - | -9.200340 | 8.175184 | - | -8.600185 | - | 5.781797 | - | - | - |
| Brain_Frontal_Cortex_BA9 | 8.769599 | 6.594851 | - | - | - | 7.463174 | - | -9.215190 | 7.449576 | 6.433872 | -8.801158 | - | - | - | - | - |
| Brain_Hippocampus | 8.548722 | 7.388470 | - | - | - | - | -6.495978 | -9.157651 | - | - | -8.370329 | - | 8.734676 | - | - | - |
| Brain_Hypothalamus | 11.412795 | 7.787702 | - | - | - | 8.567151 | - | -9.231757 | 8.339620 | - | -5.634188 | - | 6.574690 | - | - | - |
| Brain_Nucleus_accumbens_basal_ganglia | - | 10.676323 | - | -9.975992 | - | 9.210190 | - | -9.209289 | 7.451053 | - | - | - | 7.115560 | 7.332139 | - | 5.595877 |
| Brain_Putamen_basal_ganglia | - | 8.527109 | - | - | - | - | - | -9.191287 | 6.521591 | - | -8.826649 | - | 8.998337 | - | - | 5.469942 |
| Brain_Spinal_cord_cervical_c-1 | 9.852007 | 11.251321 | - | - | - | 8.429930 | - | -9.023353 | 7.873174 | - | -8.841851 | - | 5.079897 | - | - | - |
| Brain_Substantia_nigra | - | 5.872710 | - | - | - | - | - | -9.085172 | - | - | - | - | - | - | - | - |
| Pituitary | - | 7.269468 | - | - | 9.909219 | - | - | -9.217086 | 9.184165 | - | -8.699604 | - | - | - | - | 5.449284 |
| PEC_Prefrontal_cortex | - | - | 11.21723 | - | - | - | - | - | - | - | -6.173981 | - | - | - | - | 8.758629 |

**Supplementary Figure S7. Heatmap of the top 16 most significant genes across multiple brain tissues identified via Transcriptome-wide Structural Equation Modeling (T-SEM), including the MHC region.**

The heatmap illustrates tissue-specific gene expression associations, with the cells colored according to Z-scores, indicating the strength of these associations. Warmer colors (orange) correspond to higher positive Z-scores, indicating stronger positive associations, while cooler colors (blue) indicate negative Z-scores. Each row represents a brain region or tissue.
